# Detecting pathogenic structural variation in families with undiagnosed rare disease in a national genome project

**DOI:** 10.1101/2025.08.19.25333674

**Authors:** Prasun Dutta, Alistair T. Pagnamenta, Christelle Robert, Anthony E. F. McGuigan, Alison Ross, Edward S. Tobias, Ruth McGowan, Morad Ansari, David Baty, Jonathan Berg, Therese Bradley, Vera Cerqueira, Austin Diamond, Mihail Halachev, Anne Lampe, Alison Meynert, Caitlin Newman, Marian Thomson, Urmi Trivedi, Nicola Williams, Jing Yu, Javier Santoyo-Lopez, Zosia Miedzybrodzka, Jenny C. Taylor, Timothy J. Aitman

**Author notes:** These authors have contributed equally (Jenny C. Taylor, Timothy J. Aitman). Institute of Biomedical and Clinical Science, University of Exeter Medical School, Royal Devon University Healthcare NHS Foundation Trust, Exeter, Devon, UK. Sequoia Genetics, I-HUB, 84 Wood Ln, London, UK.

## Abstract

**Background:** Whole-genome sequencing (WGS) projects for rare disease diagnosis typically yield a diagnostic rate of approximately 25-40%, dependent particularly on patient selection and the extent of prior genetic testing. The Scottish Genomes Partnership (SGP) is a collaborative research programme involving four Scottish Regional Genetics Centres, four Scottish Medical Schools, and Genomics England’s 100,000 Genomes Project. It aims to facilitate genome sequencing and diagnosis for patients in the Scottish NHS with suspected rare Mendelian diseases. Within SGP, short-read sequencing (SRS) achieved a diagnostic rate of 23% in affected families.

**Methods:** To increase the diagnostic yield, we applied Oxford Nanopore Technologies (ONT) long-read sequencing (LRS) to a cohort of 24 SGP families (74 individuals) who remained undiagnosed after SRS. We also re-analysed previously generated SRS data to identify pathogenic structural variants (SVs). We benchmarked several existing software tools for SV detection using LRS and defined key requirements for sample processing and DNA quality. Custom SV prioritisation and bioinformatics pipelines were developed to integrate SV discovery with genotype-phenotype analysis.

**Results:** Benchmarking showed that minimap2 + cuteSV was optimal for single-sample SV discovery, while minimap2 + Sniffles2 performed best for family-based analysis. SV calling across the cohort yielded 60,022 filtered SVs spanning autosomes and sex chromosomes. Each family had between 23,024 and 25,009 SVs genome-wide (median: 23,814). A total of 392 SVs genome-wide and 8 within a disease-gene panel were prioritised across autosomal dominant/de novo, recessive, compound heterozygous, and X-linked modes, with counts varying between families. In three exemplar families, pathogenic or likely pathogenic *de novo* SVs were identified in both LRS and SRS data: one at the *DLX5/6* locus, one in *AUTS2*, and one in *FN1*. We provide genome-wide de novo SVs and compound heterozygous (SV + SNV) variants, and deposit raw and processed sequencing data for all families in the Genomics England Research Environment to support future gene discovery.

**Conclusions:** This study demonstrates that in-depth SV analysis can increase molecular diagnostic rates in rare disease patients with presumed monogenic aetiology. Pathogenic or likely pathogenic *de novo* SVs were identified in three families, resolving the diagnostic odyssey for at least two of the 24 families.

## Background

Rare diseases are commonly regarded as those affecting fewer than 1 in 2000 people in a population [1, 2]. Overall, up to 10,000 identifiable rare diseases have been described [1], affecting 3-6% of the population worldwide [2]. Many affected individuals remain without a molecular diagnosis, often because of nonspecific clinical presentation, resulting in long diagnostic odyssey and only symptomatic or palliative treatment [3]. Patients and carers may face psychological and emotional stress and financial difficulties in addition to the direct functional challenges of the condition. Early and accurate diagnosis can guide patient management, identify targeted treatments, and facilitate the acquisition of community-based support [4].

Depending on patient selection and prior investigations, the diagnostic rates of genome sequencing in large, rare disease cohorts range from 25% to 41% [5–9]. Traditional investigations involving karyotyping, array comparative genome hybridisation and gene- or panel-based sequencing [10–12] are now frequently complemented or replaced by whole-exome or whole-genome sequencing (WES or WGS) [13–15]. However, even with WES and WGS, most molecular diagnoses are reached from single nucleotide variants (SNVs) or small insertion□deletion (indel) variants in the protein-coding genome [6, 8, 16–18].

The most widely applied whole-genome approaches involve massively parallel short-read sequencing (SRS). While this methodology effectively covers most of the nonrepetitive genome, current knowledge limits the extent of diagnoses outside the protein-coding genome, and the most widely used analytical algorithms have limited ability to identify pathogenic variants arising from larger indels or more complex structural genomic rearrangements.

Structural variants (SVs) may be defined as those that span ≥50 bp and include genomic insertions, deletions, duplications, inversions and translocations [19]. SVs can have clinical relevance as the cause of a wide range of monogenic Mendelian and other more complex disorders [20–22]. Traditionally, structural variation has been detected by karyotyping or through imbalance being detected via array methodology. More recently, SVs have been detected via software developed to read large genomic variation signatures from SRS data [23, 24]. However, short reads suffer from mapping ambiguity, particularly in repetitive regions and problematic or low-complexity regions (segmental duplications, GC-rich regions) and often lack sensitivity for SV detection [25–27].

Long-read sequencing (LRS) technologies (or third-generation sequencing), such as those developed by Oxford Nanopore Technologies (ONT) and PacBio, have the ability to generate reads of thousands of bases up to several million bases, with the longest contiguous read of 4.2 Mb using nanopores [27, 28]. Long reads can span difficult-to-sequence regions in the genome, unambiguously providing better mappability in repetitive regions, and can span large SVs and identify split reads, pointing to inversions, deletions, duplications, or translocations that may be missed by SRS [26, 29, 30]. It is therefore possible that potentially pathogenic SVs may remain undetected in presently undiagnosed cases that have been analysed only by traditional SRS approaches [31, 32]. Although further development is still needed to refine analytical methods and reduce costs, reports of pathogenic SV identification from LRS in previously undiagnosed cases encourage the use of LRS to increase clinical diagnostic rates [31, 33–38].

The Scottish Genomes Partnership (SGP) [8] is a national research programme aligned with the Genomics England 100,000 Genomes Project [6] that aims to investigate and diagnose Scottish patients with rare Mendelian disorders via *in silico* gene panel-based analysis of short-read genome sequencing data. In SGP, SRS of 999 genomes was carried out from 394 families, each containing one or more rare disease cases of presumed monogenic aetiology. Most participants were trios, but singletons, duos and quads were also included. Sequencing data were analysed for potentially pathogenic SNVs and small indels through the Genomics England gene panel-based clinical bioinformatics pipeline, leading to an overall diagnostic yield of 23% across a range of rare disease presentations [8].

To investigate the possibility that the diagnostic rate could be increased by the identification of large structural rearrangements through LRS, 24 families who remained without a diagnosis from SGP SRS were selected for whole-genome LRS and SV analysis in the SGP programme.

Although several comprehensive studies have previously benchmarked LRS SV calling software [39, 40], they have focused primarily on single-sample SV detection. Additionally, the rapid evolution of LRS sequencing technologies and chemistries has led to continuous advancements in analytical software. Considering these factors, we conducted our own benchmarking to select the optimal SV calling tools for the SGP study. Since our primary objective was to analyse SVs in families, we focused on two SV calling software programs, CuteSV [41] and Sniffles2 [42], which support both single-sample and family-based SV calling. We also evaluated their performance with common LRS aligners.

In this study, we describe our process for SV detection and software benchmarking for choosing optimal software for our single- and family-based SV analysis from long-read sequences in SGP, which is carried out in parallel with additional analyses of whole-genome short-read sequences generated previously in the SGP programme. We report exemplar cases of pathogenic or likely pathogenic SVs identified with these approaches.

## Methods

### Patient Recruitment and Sample Collection

#### Recruitment criteria for SGP LRS

Twenty-four families recruited for the SGP LRS study were selected from those that did not receive a genetic diagnosis in the SGP SRS study [8]. The Scottish families recruited for SRS within the SGP presented a rare phenotype that was believed to be genetic in origin. Families had variable structures (singletons, duos, trios, and quads) and had negative results from prior diagnostic testing, including microarray analysis.

To be selected for SGP LRS, families needed to be trios or quads within the SGP SRS study, with a negative report following both primary assessment (Tiers 1 and 2 variants) and secondary assessment (Exomiser [43]/in-depth investigation of Tier 3 variants) [8]. The selected families had a high index of suspicion of monogenic aetiology, either because multiple siblings or family members had the same phenotype or because of syndromic presentation suggestive of monogenic disease. The focus was therefore on families that might benefit the most from diagnosis through LRS analysis. In addition to the 24 families (comprising 74 participants), an additional 4 participants from the same cohort were included as part of a pilot study for sequencing platform optimisation.

#### DNA preparation

Details of sample collection and DNA extraction were the same as those used for the SGP SRS study [8]. Briefly, DNA was extracted from blood at various Scottish Regional Genetics Laboratories (Aberdeen, Dundee, Edinburgh, and Glasgow). To be selected for the SGP LRS study, a minimum of 15 µg of genomic DNA, preferably with a DNA integrity number (DIN) value of > 8.0, was required to be available from all family members.

### Sample processing and sequencing

#### Genomic DNA fragmentation and size selection

Approximately 7.5 µg of high-molecular-weight (HMW) genomic DNA was sheared into fragments of ∼25--40 kb via Megaruptor 3 (Diagenode) according to the Megaruptor instructions at speed 28. A 1x volume of AMPure XP beads was added to the sheared material, and the supernatant was discarded after 30 minutes at room temperature. The beads were washed twice with 80% ethanol, and the DNA was eluted in 60 µl of TE after incubation at 37°C for 1 hour. The fragment size was estimated via pulsed-field capillary electrophoresis via the Femto Pulse system (Agilent Technologies, Santa Clara, CA, USA). To remove short DNA fragments, 30 µl of sheared DNA was loaded into a well of a BluePippin (Sage Science, Beverly, MA, USA) agarose gel cassette (0.75% DF) with 10 µl of loading solution. After 6 hours, 20-40 kb DNA fragments were collected, following the high-pass DNA size selection instructions using marker S1. A total of 40 µl of eluate was transferred to a fresh tube, and 1x volume AMPure XP beads was added. The beads were incubated for 30 minutes at room temperature, and two washes with 80% ethanol were performed. The DNA was eluted in 50 µl of nuclease-free water after 1 hour of incubation at 37°C. DNA was quantified via the Qubit dsDNA High Sensitivity Assay (Thermo Scientific, Wilmington, NC, USA).

#### Library preparation for Oxford Nanopore genome sequencing

For the pilot study, four participants were selected from the SGP SRS study. For the first three participants, two libraries were constructed, where shearing and size selection were not performed in the first library, whereas shearing and size selection were performed in the second library. Shearing and size selection resulted in the concentration of DNA fragments with lengths between 10 and 40 kb. A sample of partially degraded genomic DNA was available from the fourth selected participant, with a low proportion of high-molecular-weight DNA (less than 40% of the gDNA was >40 kb). Size selection was also performed on this sample, with no further shearing. This led to a total of 7 libraries being sequenced on the ONT PromethION Beta machine for the pilot study. The same library preparation and sequencing method was used for family-based samples (n=74).

For the substantive family study, 73 samples were sequenced on ONT R9.4.1 flow cells via a V9 ligation sequencing kit (SQK-LSK109, Oxford Nanopore Technologies, UK). This included the four participants from the pilot study, who were resequenced as part of the expanded cohort. A further 28 of these samples with adequate residual DNA were resequenced via the R10.4.1 (FLO-PRO114M) flow cell via the V14 ligation sequencing kit (SQK-LSK114, Oxford Nanopore Technologies, UK) for library preparation. This was done to achieve greater depth of coverage utilising the higher yield property of the R10.4.1 ONT flow cells. One additional sample was sequenced only on an R10.4.1 flow cell, resulting in a total of 74 samples sequenced for the family-based SV study (Additional File 1: Supplementary Table 1). No size selection was performed on samples sequenced on the R10.4.1 flow cells.

Approximately 1-2 µg of sheared and size-selected DNA in a volume of 47 µl of nuclease-free water was used with the SQK-LSK109 or SQK-LSK114 ligation sequencing kits (Oxford Nanopore Technologies, UK), depending on the flow cell type used, for library preparation following Oxford Nanopore recommendations. Individual sample preps were loaded into a single R9.4.1 and run on a PromethION Beta (Oxford Nanopore Technologies, UK) for 72 hours or a single R10.4.1 flow cell and run on the PromethION 24 (Oxford Nanopore Technologies, UK) for 90 hours. Sequencing was carried out at the Edinburgh Genomics facility, University of Edinburgh, UK.

### SV analysis

#### Software benchmarking

Three long-read-based aligners and two long-read-based SV callers were chosen for benchmarking. For alignment, minimap2 [44] v2.23-r1111, NGMLR [25] v0.2.7 and lra [45] v1.3.2 were chosen because of their ability to align long reads. For SV detection, cuteSV v1.0.13 and Sniffles v2.0.2 were chosen. The aim of the benchmarking process was to find the best aligner and SV caller combination suitable for accurate detection of potentially pathogenic SVs. The latest available versions of each software were used at the time of analysis.

For benchmarking, the truvari v3.1.0 [46] *bench* program was used to calculate the recall (true-positive rate), precision and F1 score, or F-measure (accuracy). Benchmarking was performed on Ashkenazi trio Son (Genome in a bottle [47] or GIAB HG002) Oxford Nanopore data. The truth set consisted of 5,262 insertions and 4,095 deletions [48] >=50 bp. Alignment and SV calling were performed against the GRCh37 build of the human reference genome, as the truth set belonged to that version.

SV calling was performed via the two SV callers (CuteSV and Sniffles2) on each BAM file, resulting in two VCF files from each aligner-SV caller combination (a total of 6 VCFs). A consensus VCF was created via SURVIVOR [49] *merge* v1.0.7, which contained SVs that were common to any four VCF files, had the same SV type, were present in the same strand, and had a maximum distance between the breakpoints of 500 bp. Since the truth set consists of only insertions and deletions, all VCFs were pre-processed by a custom script where all duplications were converted to insertions and only insertions and deletions were retained.

To infer the optimal filtering thresholds of two variant metrics—GQ (Phred-scaled genotype quality) and QUAL (Phred-scaled variant quality)—output from the truvari program was utilised. For GQ, the --gtcomp parameter was used to select only those SVs whose genotypes matched with the truth set. For testing QUAL, this parameter was avoided.

#### SV analysis and filtering

In the pilot study (n=7), base calling and sample demultiplexing were performed via Guppy (Oxford Nanopore Technologies, UK) v4.0.11+f1071ce. The resulting reads that passed Guppy QC were put into a FASTQ file with a “pass” tag. Further QC was performed in this FASTQ file in the same way as the benchmarking process to create trimmed and filtered reads. FASTQ statistics for the 7 samples were generated via NanoComp v1.17.0.

The alignment of trimmed and filtered FASTQ files was performed via minimap2 v2.23-r1111 against the GRCh38 build of the human reference genome via the parameters -x map-ont and -MD. SVs were called via cuteSV v1.0.13. BAM file statistics were calculated via Qualimap [50] *bamqc* v2.2.2-dev and NanoComp [51] v1.17.0.

In the family-based study (n=74), all samples were sequenced via the R9.4.1 flow cell on the PromethION Beta, and 29 additional samples were sequenced via the R10.4.1 flow cells on the PromethION 24 machine (Additional File 1: Supplementary Table 1). For base calling and demultiplexing, Guppy v5.1.13+b292f4d was used in the PromethION Beta machine, whereas Guppy v6.5.7+ca6d6af was used in the PromethION 24 machine. QC on the resulting FASTQ files with the “pass” tag was performed in the same manner as above, except that reads below 1000 bp were discarded. Read statistics were calculated in the same way as above.

Trimmed and filtered reads (FASTQ) were aligned to the GRCh38 reference genome via minimap2 v2.23-r1111 with parameters -x map-ont, --MD and --secondary=no and coordinate-sorted and indexed via samtools [52, 53] v1.15.1. The FASTQ files generated from each flow cell type were processed separately to produce trimmed and filtered reads. For samples that were sequenced on both R9.4.1 and R10.4.1 flow cells, BAM files were merged into a single combined BAM file. QC statistics were calculated on the resulting BAM files via NanoComp v1.17.0, Qualimap *bamqc* v2.2.2 and samtools *flagstat* v1.15.1. Multisample SV calling was performed across all SGP LRS samples (n=74) via Sniffles v2.0.6, where first, snf (Sniffles2 file, a specialised binary file format introduced in [42]) files were produced from the BAM files via --long-del-length 1000000, --long-ins-length 1000000, --long-dup-length 1000000, --minsvlen 50 and --tandem-repeats, and then multisample SV calling was performed via all 74 snf files, producing a fully genotyped multisample VCF file. SVs overlapping with genome gaps and contamination/false duplication were removed. Information on gaps in regions such as short_arm, heterochromatin, telomere, contig and scaffold regions was downloaded from UCSC for hg38, and any overlapping SVs were removed. Monomorphic loci and any loci with a >90% missing data rate were also removed. Those SVs that had average site mean depth (or mean depth per site averaged across all individuals) < 10 and > 56 were removed. This removed SVs that are present/detected in places that have extremely high depths of coverage due to alignment artifacts/repeats, etc. The maximum site mean depth is dependent on the mean depth across all SV sites and is double that value. SVs with a QUAL < 25 were removed. Furthermore, SVs with lengths of 50 bp or more and missing lengths (to account for translocations) were retained. All filtering was performed via BCFtools v1.15.1 and bedtools v2.30.0. Finally, SV annotation and functional impact prediction were performed via VEP v110.1, and panel gene-specific SVs were selected via a VEP *filter* in a custom script. SV allele frequency annotation was performed via SVAFotate [54] v0.1.0 (parameters: -f 0.5, --cov 0, --lim, 1000000, -a mis), which annotates SVs via population-level allele frequency (AF) values from four major databases: CCDG [55], gnomAD-SV [21] (v4.2.1), 1000G [56] and TopMed SVs [57]. SVAFotate annotates each SV with the maximum AF (Max_AF) from all matching SVs across all four databases. In this study, SVs are denoted as common if Max_AF >= 0.05, low frequency if Max_AF < 0.05 and Max_AF >= 0.01 and rare if Max_AF < 0.01. If Max_AF = 0, then the SV is unique to the SGP LRS cohort. Notably, the unique SV might have a different SV type in the population databases. The complete SV analysis and filtering schematic diagram is shown in Additional File 1: Supplementary Figure 1.

#### SV prioritisation

To prioritise *de novo* SVs, the multisample VCF file with autosomal SVs was processed via BCFtools v1.15.1 to obtain family-specific VCF files (n=24), after which monomorphic loci were removed. Slivar [58] v0.3.1 was used to find *de novo* SVs in the family VCFs, which followed this rule: the proband’s genotype is a heterozygous alternate or homozygous alternate, the mother and father have a homozygous reference genotype (0/0 for mother and father and 0/1 or 1/1 for proband), the proband, mother and father should have a minimum depth of 5 reads, and there should be no reads in the mother and father supporting the variant/alternate allele, Max_AF < 0.001 and SGP LRS cohort AF < 0.01. Additionally, SGP LRS cohort AF < 0.015 filtering was also performed separately (keeping all other filtering parameters the same) to detect *de novo* SVs that could have been genotyped incorrectly.

Patient-specific clinician-assigned gene panels were downloaded from PanelApp [59]. The latest versions of the diagnostic-grade green label gene panels were considered for this analysis. Only those *de novo* SVs that overlapped with genes from panels assigned to the family by their respective clinicians were retained. Only panel genes that showed a monoallelic mode of inheritance (MOI) linked to the disorder represented were retained via a custom script.

To identify SV overlapping genes with homozygous autosomal recessive MOIs for the condition in question, genes annotated with biallelic MOIs were selected from panels associated with the proband. Those SVs were prioritised via slivar where a proband showed a homozygous genotype for the alternate allele and where the mother and father were heterozygous for the alternate allele (0/1 for the mother and father and 1/1 for the proband), all three samples had a depth of at least 5 reads and Max_AF < 0.01 and the SGP LRS cohort AF < 0.05.

Potential compound heterozygotes with an SV in-*trans* with an SNV or small indel were also investigated. Previously generated Illumina short-read data [8] were used to detect SNVs and small indels because of their greater depth, whereas SVs detected from LRS were used for this analysis. Short-read Illumina VCF files with small variants (SNVs and indels) created by Edinburgh Genomics were intersected with long-read structural variant (SV) VCFs. On a per-family basis, proband SVs were retained where they passed all quality filters, the SGP LRS cohort AF was < 0.01, the biotype was “protein_coding”, IMPACT was HIGH or MODERATE, and the overlapping transcript was canonical. The corresponding SNV VCF files were then generated via this subset of regions that passed all the quality filters. The SNV VCFs were then normalised via BCFtools v1.20 to decompose multiallelic sites and then annotated with VEP v110, including annotations for precomputed ‘masked’ spliceAI [60] scores, CADD [61] scores and gnomAD [62] v4.1 allele frequency annotations. SNVs were filtered using gnomAD AF (exomes + genomes) < 0.01, and an IMPACT was either HIGH or MODERATE. A compound heterozygote was considered when at least one SV and SNV passed this filtering with an inheritance pattern suggesting that the variants were in-*trans*. Panel-specific compound heterozygous scenarios were then obtained as a subset of the genome-wide scenarios, focusing on biallelic panel genes.

For X-linked analysis, those families that had an affected male proband with a hemizygous SV and an unaffected/carrier mother with a heterozygous SV were examined; Max_AF < 0.01, the SGP LRS cohort AF < 0.05, and all samples of the family had a depth of at least 5 reads. Both panel-based and chromosome-wide analyses were carried out.

#### SRS SV analysis using SVRare

Genome sequencing and data processing in the 100,000 Genomes Project have been described elsewhere [6]. In brief, libraries were prepared from DNA extracted from blood via the TruSeq PCR-Free High-Throughput Kit, and 150 bp paired-end reads were sequenced in a single lane of an Illumina HiSeqX instrument. The reads were aligned to the GRCh38 build or GRCh37 build of the human reference genome via the iSAAC Aligner [63], and the SVs were called via Manta [64] and Canvas [65].

SVRare [66] was used to collate the deletions, duplications and inversions of the SVs into a database and annotate each SV with an estimated recurrence frequency, the potential to disrupt the protein-coding sequence of the overlapping genes, and its relevance to the patient’s disease or phenotypes. SVs with GRCh37 coordinates were lifted over to GRCh38, and an 80% overlap threshold was used for recurrence frequency estimation.

### Quality Control

#### Mendelian consistency analysis

A Mendelian consistency analysis was performed in one of the trios from the SGP LRS cohort, where the family was analysed for the presence of Mendelian errors that may be candidates for *de novo* mutation. This analysis was also performed elsewhere [42] on the Ashkenazi Jewish trio. The analysis could also indicate which SV-calling software is most suitable for family-based SV analysis in the SGP LRS cohort.

Minimap2 v2.23-r1111-aligned BAM files were subjected to force-calling (two-pass mode) via cuteSV v1.0.13, force-calling via Sniffles v2.0.6 and a multisample (family-based) SV calling procedure via Sniffles v2.0.6. In force calling via cuteSV, SVs are first called individually. Only precise SVs with QUAL > 20 and GQ > 15 were retained. The filtered VCFs were then merged via SURVIVOR to create an integrated/joint VCF file. This merged VCF file was then used to regenerate individual VCFs again. The regenotyped VCFs were again filtered to retain precise SVs with QUAL > 20 and GQ > 15. The filtered VCFs were then merged again via SURVIVOR to obtain the final trio VCF file. Sniffle force calling was performed in the same manner, but Sniffles2 was used for calling SVs. For Sniffles v2.0.6 multisample calling VCF, first, Sniffles binary files were produced from individual BAM files from the trio, which were then used jointly to produce a trio VCF file. Only those SVs that were precise and had a QUAL > 20 were retained, and the genotypes were classified as missing (./.) where GQ < 15. BCFtools [53, 67] +Mendelian v1.15.1 plugin with the -m c parameter (print counts) was used to calculate statistics on Mendelian consistency on the final trio VCF files from all three pipelines.

#### SV read length analysis

An SV read length analysis was carried out on the pilot study samples. For this analysis, only those participants who had both size-selected and non-selected libraries were selected (n=3, pilot study). The BAM files generated from size-selected and non-selected libraries that belonged to the same participants were merged, and SVs were called via cuteSV v1.0.10, resulting in three VCF files. The three VCF files were combined via SURVIVOR *merge* v1.0.7 to obtain a union set of SVs. Those SVs were merged where the maximum distance between the breakpoints was 500 bp, were called by cuteSV in at least one VCF, had the same SV type, were present in the same strand and were at least 50 bp in length. The SV length distribution of the union set was plotted via surpyvor *lengthplot* v0.12.0 with the -all parameter. All the SVs present in the VCF file were plotted regardless of zygosity. Additionally, to account for all the SVs discovered, the ‘plots.py’ program of surpyvor was edited to accommodate SVs up to 100 kb. By default, surpyvor *lengthplot* plots SVs only up to 20 kb.

## Results

### Benchmarking results for single-sample SV calling

For single-sample benchmarking and SV calling, SV calls in the son of the Ashkenazi Jewish trio were evaluated via three long-read-based aligners (minimap2, NGMLR and lra) and two SV callers (cuteSV and sniffles2). The mean depth of coverage was 33x across the human genome and was the same for all three aligners. Table 1 shows the benchmarking results in terms of precision and recall (sensitivity). The highest recall was achieved by the minimap2_cuteSV combination (0.93), whereas the highest precision was achieved by the consensus caller (consensus_caller_4_set) and lra_sniffles combination (0.98). However, lra_sniffles had the lowest recall rate of 0.69. The highest F-measure (F1 score) was shown by minimap2 alignment and cuteSV SV calling (0.93).

**Table 1:** Performance metrics for various aligner and SV-caller combinations. Table 1: Precision, recall and F1 score statistics for different aligner and SV-caller combinations for different mean depths of coverage (33x and 10x subsampled).

| Mean Depth of coverage | 33x |  |  | 10x |  |  |
| --- | --- | --- | --- | --- | --- | --- |
| aligner_SV-caller | Precision | Recall | F1 | Precision | Recall | F1 |
| minimap2_cuteSV | 0.94 | 0.93 | 0.93 | 0.95 | 0.67 | 0.79 |
| ngmlr_cuteSV | 0.92 | 0.92 | 0.92 | 0.94 | 0.66 | 0.77 |
| lra_cuteSV | 0.95 | 0.88 | 0.91 | 0.96 | 0.61 | 0.75 |
| minimap2_sniffles | 0.97 | 0.77 | 0.86 | 0.97 | 0.55 | 0.71 |
| ngmlr_sniffles | 0.96 | 0.75 | 0.84 | 0.97 | 0.53 | 0.69 |
| lra_sniffles | 0.98 | 0.69 | 0.81 | 0.98 | 0.48 | 0.64 |
| consensus_caller_4_set | 0.98 | 0.81 | 0.89 | 0.98 | 0.57 | 0.72 |

For the subsampled set derived from the Ashkenazi Jewish trio son (mean depth of coverage of 10x), the consensus caller and lra_sniffles combination showed the highest precision of 0.98 (Table 1). In terms of recall, the lra_sniffles combination had the lowest value (0.48), and minimap2_cuteSV had the highest value (0.67). In terms of the F1 score, minimap2_cuteSV was at the top (0.79).

When only those SVs whose genotypes were the same as those of the SV type were considered, the minimap2_cuteSV combination still presented the highest F1 score for both 33x and 10x mean depths of coverage (Table 2).

**Table 2:** Performance metrics of genotype (GT) concordant SVs for different aligner and SV-caller combination. Table 2: Precision, recall and F1 score statistics of genotype (GT) concordant SVs for different aligner and SV-caller combinations for different mean depths of coverage (33x and subsampled 10x). The values are calculated for SVs whose genotypes match the genotypes of the SVs in the truth set.

| Mean Depth of coverage | 33x |  |  | 10x |  |  |
| --- | --- | --- | --- | --- | --- | --- |
| aligner_SV-caller | Precision-GT | Recall-GT | F1-GT | Precision-GT | Recall-GT | F1-GT |
| minimap2_cuteSV | 0.92 | 0.91 | 0.92 | 0.88 | 0.62 | 0.73 |
| ngmlr_cuteSV | 0.90 | 0.90 | 0.90 | 0.87 | 0.60 | 0.71 |
| lra_cuteSV | 0.93 | 0.85 | 0.89 | 0.89 | 0.56 | 0.69 |
| minimap2_sniffles | 0.96 | 0.76 | 0.85 | 0.93 | 0.53 | 0.67 |
| ngmlr_sniffles | 0.90 | 0.70 | 0.79 | 0.87 | 0.48 | 0.62 |
| lra_sniffles | 0.96 | 0.67 | 0.79 | 0.93 | 0.46 | 0.61 |

### Family-based SV calling and Mendelian inconsistency analysis

Family-based variant calling using parents and offspring allows prioritisation of variants on the basis of the MOI of genes/disease, may help reduce the number of variants to be analysed and can accurately identify *de novo* variants that cannot be identified if only the affected individual is sequenced [16].

We observed that Sniffles2 multisample/family calling had the highest number of genotypes with no missingness (i.e., absence of genotype calls at a given variant position) and Mendelian errors (%OK or consistent) of approximately 59% (Additional File 1: Supplementary Figure 2). Both cuteSV force calling and Sniffles2 force calling had lower %OK values. The number of Mendelian errors was highest for Sniffles2 family calling, whereas % skipped or the number of genotypes skipped due to missingness was lowest in Sniffles2 family calling. Based on these results, Sniffles2 was chosen to call SVs in the SGP LRS samples.

### Hard-filtering threshold analysis for single-sample SV calling

To reduce false positive or artifactual SVs after SV calling, we conducted a hard-filtering threshold analysis on genotype and variant quality metrics to determine a suitable threshold that helps retain true positive SVs. This analysis was performed on the Ashkenazi Jewish trio son. According to earlier benchmarking tests, for single-sample SV calling, minimap2 as an aligner and cuteSV as the SV caller demonstrated the highest performance in terms of the F1 score (Table 1). Two metrics/annotations were chosen based on which high-quality SVs could be filtered, GQ and QUAL, with the distributions shown in Figure 1a and b. The subsampled BAM (10x mean depth of coverage of 10x of the Ashkenazi Jewish triple son sample) was used to find the SVs for threshold analysis after alignment via minimap2 and SV calling via cuteSV. The aim of this analysis was to find an appropriate threshold for the chosen metrics that would allow us to keep the highest number of true-positive (TP) SVs and reduce the number of false-positive (FP) SVs. Figure 1a shows the distribution profile of GQ values for TP SVs and FP SVs. Most of the TP SVs had GQ ≥ 20, and the distribution peaked at 20. Furthermore, it was observed that most of the FP SVs had GQ ≤ 10. There is an overlap between the two distributions, and they intersect at GQ 15. Based on this plot, it was inferred that GQ=15 can be chosen as an appropriate threshold for GQ.

**Figure 1:**
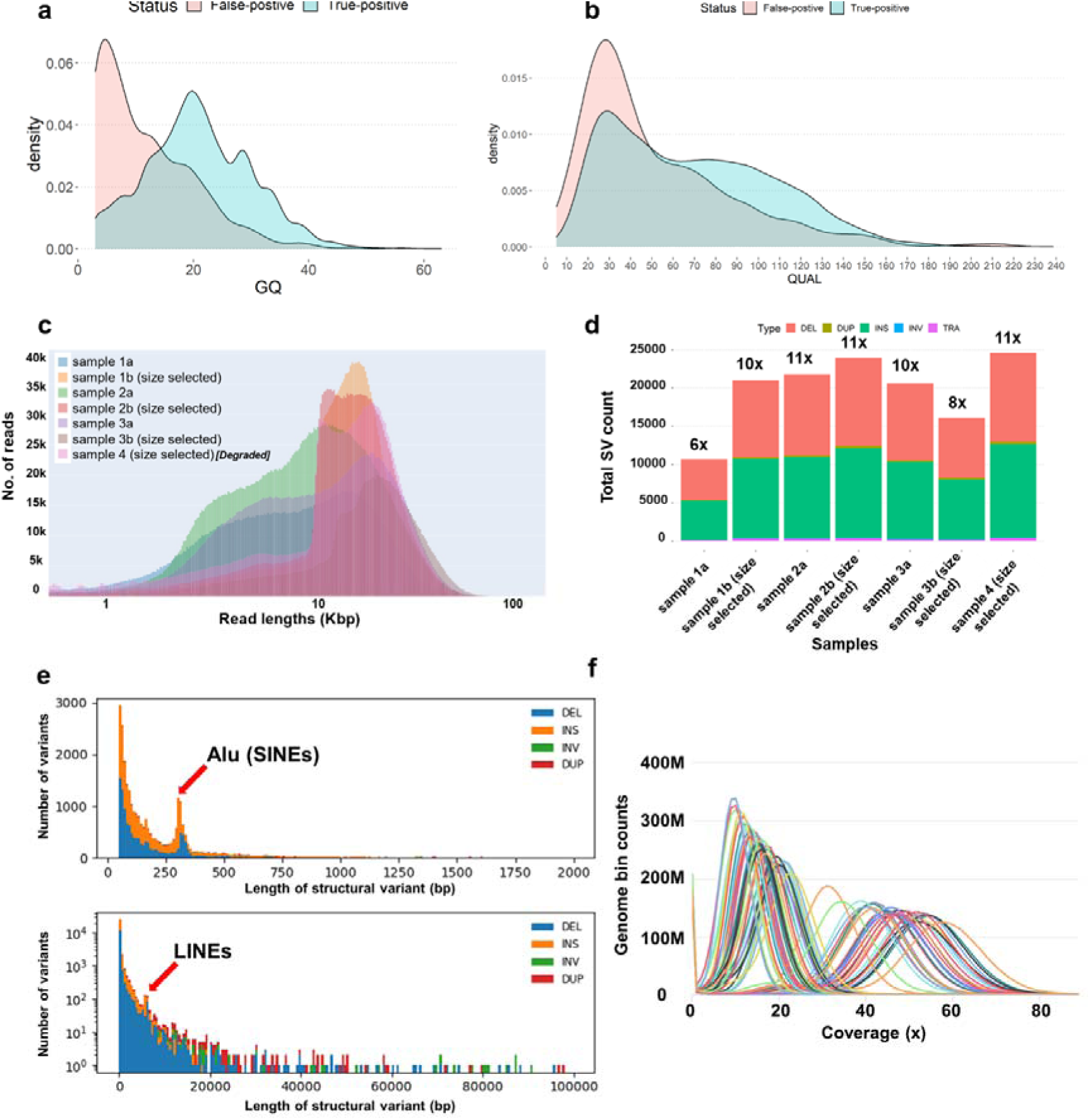
(a) Density plot for the genotype quality (GQ) score distribution for true-positive (TP) and false-positive (FP) SVs. (b) Density plot for variant quality (QUAL) for TP and FP SVs. (c) Histogram of log-transformed read length (in kb) for size-selected and non-size-selected samples in the pilot study (n=7) produced via NanoComp. Different samples are shown in different colours in the plot. (d) Histogram of total SVs discovered in each of the 7 samples. Five SV types (insertions (INS), deletions (DEL), duplications (DUP), inversions (INV), and translocations (TRA)) are denoted by different colours. Sample 4 (size selected) refers to the degraded sample. (e) SV length distribution plots plotted via survpyvor (upper panel) histogram showing SVs up to 2 kb binned per 10 bp (lower panel) histogram showing SVs up to 100 kb binned per 500 bp. The y-axis is on a log scale. Lengths of four types of SVs are plotted in both plots: deletions (DELs), insertions (INSs), inversions (INVs), and duplications (DUPs), and the length is in absolute value. Zygosity was not considered when the plots were constructed. Spikes at ∼300 bp and ∼6 kb correspond to Alu and LINE insertions, respectively. (f) Coverage histogram for SGP LRS samples (n=74) showing the distribution of the number of locations in the reference genome with a given depth of coverage. Each sample is represented by a different line color. The average depth of coverage distribution ranges from approx. 10x to approx. 50x across the 74 samples. Merged (R9.4.1+R10.4.1) BAM files were used to generate this plot.

Most of the TP SVs had a QUAL ≥ 30, and most of the FP SVs also had a QUAL ≥ 30 (Figure 1b). There is also good overlap between the TP and FP SV’s QUAL distributions. On the basis of the plot, QUAL=20 was chosen as an appropriate threshold. A value of 50 or even 30 could also have been chosen at the intersection of the distributions, but using these thresholds would result in the loss of a substantial number of TP SVs.

### Effect of size selection and DNA fragmentation on sequence outputs

This analysis aimed to check the suitability of clinical samples of varying quality and to determine the effects of size selection and DNA fragmentation on SV discovery. A total of 7 samples were included in the study. To assess sample suitability for LRS and SV discovery, we sequenced 7 singleton samples on the R9.4.1 flowcell. Read alignment against GRCh38 via minimap2 resulted in a median depth of coverage of 6--11x, whereas the mean depth of coverage ranged from 6.8x--11.5x. NanoComp and Qualimap (mean and median coverage) calculated alignment statistics are shown in Table 3.

**Table 3:** BAM file statistics from pilot study samples. Table 3: BAM file statistics for 7 pilot samples calculated via Qualimap and NanoComp. Only primary alignments are considered in read counts.

| Sample Name | Total sequenced bases (yield) | Median read length | Read length N50 | Aligned Reads | Median Coverage | Mean Coverage |
| --- | --- | --- | --- | --- | --- | --- |
| sample 1a | 20178537964 | 7494 | 14459 | 2107646 | 6.0x | 6.8x |
| sample 1b (size selected) | 31808832054 | 14074 | 17001 | 2187913 | 10.0x | 10.5x |
| sample 2a | 34145543065 | 8739 | 15046 | 3199074 | 11.0x | 11.5x |
| sample 2b (size selected) | 34863271406 | 13751 | 17757 | 2402842 | 11.0x | 11.5x |
| sample 3a | 32202547854 | 10037 | 18305 | 2609081 | 10.0x | 10.9x |
| sample 3b (size selected) | 25324502117 | 17185 | 22736 | 1429244 | 8.0x | 8.4x |
| sample 4 (size selected) | 34384235565 | 13233 | 18502 | 2488596 | 11.0x | 11.4x |

We observed that the total number of sequenced bases (or yield) was related to the median and mean coverage even though the number of aligned reads did not (Table 3). The median read length (and read length N50) is greater in sample 3b (size selected) than in the former. Longer reads can overlap with each other, increasing confidence over a position and increasing the locus-based depth of coverage. In most cases, the read length seems to be related to the coverage, except for sample 3b (size selected), where even though the median read length is the highest, it could only achieve a median coverage of 8x. On the basis of these inferences, it can be concluded that in this study, both yield and read length play important roles in achieving our desired depth of coverage.

We observed that there was no effect of size selection on mapping, as a non-sized selected sample (sample 2a) with a higher yield achieved a greater depth of coverage (median coverage of 11x) than a size-selected sample (sample 3b (size selected)), where a median coverage of 8x was achieved after mapping (Table 3). Furthermore, sample 4 (size selected), which was degraded, also performed well, with a mean and median depth of coverage ≥11x. This finding supported the possibility of using degraded samples with size selection in this study.

We observed that the size-selected samples had longer median read lengths than the non-selected samples did (Table 3). To better understand if the size selection procedure was successful, the read length distributions from the trimmed and filtered raw FASTQ reads were plotted for the seven samples in Figure 1c. The size selection protocol indeed works, as, in the size-selected samples, most of the reads were ≥10 kb in length. On the other hand, the distribution of the read lengths of the non-selected samples starts to peak at approximately 5 kb. We also observed that some of the read lengths of size-selected samples were up to 80-100 kb, with the partially degraded sample producing equally good results as nondegraded samples, with most of the read lengths being ≥10 kb.

### SV calling on pilot samples (n=7)

The pilot study samples were subjected to SV calling via cuteSV. We wanted to establish whether the samples in the study were suitable for SV discovery and, accordingly, how many SVs could be discovered from the clinical samples with respect to the reference genome (GRCh38) at a depth of coverage of 10x. For this analysis, the SVs were not subjected to any QC. Approximately 10,000-25,000 SVs could be detected across the 7 samples (Figure 1d). The depth of coverage (and yield) influenced the number of SVs discovered. ≥20,000 SVs could be detected at depths of 10x and above, and the majority were insertions and deletions, which is similar to the results achieved in recent studies [68, 69]. Duplications, inversions, and translocations were also discovered but were fewer in number than insertions and deletions were. A high number of SVs were detected in the degraded sample (sample 4 (size selected)), which indicated that the degraded clinical samples were suitable for SV discovery. Figure 1d clearly revealed that the number of SVs discovered was influenced by and correlated with the mean depth of coverage.

### SV length distribution in pilot samples (n=7)

The union set of SVs generated (SVs present in at least one VCF) resulted in approximately 32,000 SVs after retaining SVs ≥ 50 bp. In Figure 1e (upper panel), at approx. 300 bp, a peak is observed for insertions and deletions, and they refer to the *Alu* mobile transposable element, which is classified as short interspersed nuclear elements or SINEs. A second peak of insertions and deletions is observed at approximately 6000 bp in Figure 1e (lower panel), and they refer to L1 or long interspersed nuclear elements (LINE-1). This finding is consistent with those of previous studies [48, 70]. Figure 1e (lower panel) shows that SVs ≥ 80 kb in length were detected.

### Sequencing results of family samples (n=74)

For the family-based study, 74 individuals were sequenced in total. They belong to 24 families and consist of 22 trios and 2 quads (Additional File 1: Supplementary Table 1). In total, 6.71 Tb of sequencing data were generated by the ONT PromethION Beta and PromethION 24 machines, where 45 samples were sequenced via only R9.4.1 flow cells and 29 samples were sequenced via both R9.4.1 and R10.4.1 flow cells. For the raw data sequenced via the R9.4.1 flow cell, the number of reads per sample (n=73) ranged from 1,973,096 to 5,594,691 (Additional File 1: Supplementary Table 2). The trimmed and filtered statistics for n=73 samples are presented in Additional File 1: Supplementary Table 3. For samples sequenced via R10.4.1 flow cells (n=29), the number of reads ranged from 11,369,179 to 53,919,664 (Additional File 1: Supplementary Table 4). The trimmed and filtered statistics for n=29 samples are shown in Additional File 1: Supplementary Table 5.

To visualise the distribution of key QC metrics across different samples (for samples sequenced in R9.4.1 (n=73) and R10.4.1 flow cells (n=29), four metrics were plotted: median read length, median read quality, number of reads and read length N50 (Additional File 1: Supplementary Figure 3). In both sets, when filtering was applied to the raw reads, it led to increases in the median read length, median read quality and read length N50 (Additional File 1: Supplementary Figure 3 a and c). This was due to the removal of short reads, phage sequences and low-quality bases in both sample sets. A clear result postfiltering was that the number of reads decreased when reads of lower quality were removed (also reflected by the increase in median read quality between the raw and filtered samples). The percentage difference between the raw and filtered samples for each metric is shown in Additional File 1: Supplementary Figure 3 b and d. The median read quality for samples sequenced via the R10.4.1 flow cells was greater than that for samples sequenced via the R9.4.1 flow cells and had a tight distribution, indicating that the former produced reads of consistent quality. A much greater number of reads (signifying flow cell yield) was produced by the R10.4.1 flow cells than by the R9.4.1 flow cells. An additional analysis of the flow cell yield and depth of coverage is shown below. Filtering the raw data from the samples sequenced via the R9.4.1 flow cells led to a difference of approximately 10%. However, for samples sequenced via the R10.4.1 flow cells, there was a difference of approximately 40%. This might have occurred since no size selection was performed on these samples, and many shorter reads were sequenced, which were eventually removed after filtering. There is also a marked difference between the median read lengths of the two flow cells. The samples sequenced via the R10.4.1 flow cells presented greater standard deviations in the read length distribution (data not shown), indicating high variability in read length.

Since two flowcell versions were used in our study, we compared the performance of the two flowcell versions. From the depth of coverage plots for samples sequenced using only R9.4.1 (73 samples) and only R10.4.1 flow cells (29 samples), it is clearly observed that the latter’s greater yield led to greater depth (Additional File 1: Supplementary Figure 4). This is important, as greater depth helps confidently genotype variants downstream and prevents sampling bias. A greater depth was further achieved by combining the data from both flow cells. The distribution of the depth of coverage across the 74 samples is shown as a coverage histogram in Figure 1f. The alignment of trimmed and filtered reads to the human reference genome (GRCh38) led to a mean depth of coverage of 10.1x to 57.2x. Across the 74 samples, ≥ 99% of the total reads successfully mapped to the human genome, with an average percent identity (% of nucleotides that match with the reference genome) of 91.50--97.20%. The complete statistics of the BAM files are shown in Additional File 1: Supplementary Table 6. Notably, the number of total reads is greater in the BAM file statistics than in the total reads in trimmed and filtered FASTQ file statistics because the former contains additional supplementary read counts that signify split reads that the aligner (minimap2) creates during alignment, for example, in the presence of an SV.

### Overall SV-calling statistics in SGP LRS families

Multisample SV-calling resulted in the discovery of 74,388 SVs across 74 individuals. Regions with an unusually high depth of coverage resulting from read mapping artefacts or due to repetitive regions of the genome might lead to spurious SV calls. To detect such SVs, the distribution of the mean depth of the SVs (site mean read depth) was visualised (Figure 2a). Most of the SVs in our dataset had an approximately 28x mean depth of coverage (averaged across all samples), as seen from the plot. Therefore, a minimum of 10x mean depth was chosen to remove SVs with low mean read depth, and a maximum of 56x (28*2) was chosen to remove SVs with unusual depth of coverage. After performing read depth filtering and other filtering procedures (see Methods), a total of 60,022 SVs remained across autosomes and sex chromosomes. Most of the discovered SVs (61.9%) were unique to the SGP LRS family (Max_AF = 0), and no matches were found by SVAFotate in its databases (Additional File 1: Supplementary Figure 5 and Figure 2b), followed by common (21.33%), rare (10.9%) and less frequent (5.87%). Figure 2e shows the percentage distribution of SVs being common, less frequent, rare and unique when stratified by SV type and followed the same trend as that in the overall dataset, with the highest number of SVs being unique except for duplications, where rare SVs were higher in number. As shown in Figure 2c, the highest number of SVs in the SGP LRS cohort were insertions and deletions (52% and 44.33%, respectively), followed by translocations/BND (3.1%), duplications (0.3%) and inversions (0.3%). A total of 23,024-25,009 SVs were called across 24 families (median: 23,814) (Figure 2d), of which 12,416-13,489 SVs were insertions, 10,087-11,080 SVs were deletions, 13-37 SVs were duplications, 33-57 SVs were inversions, and 361-679 SVs were translocations. The distribution of the number of different SV types discovered in the 24 families did not show any major outliers (Figure 2f and g). The SV statistics are shown in Additional File 1: Supplementary Table 7, and the samples used to derive this table consist of both R9.4.1 and R9.4.1+R10.4.1 data (when both are available).

**Figure 2:**
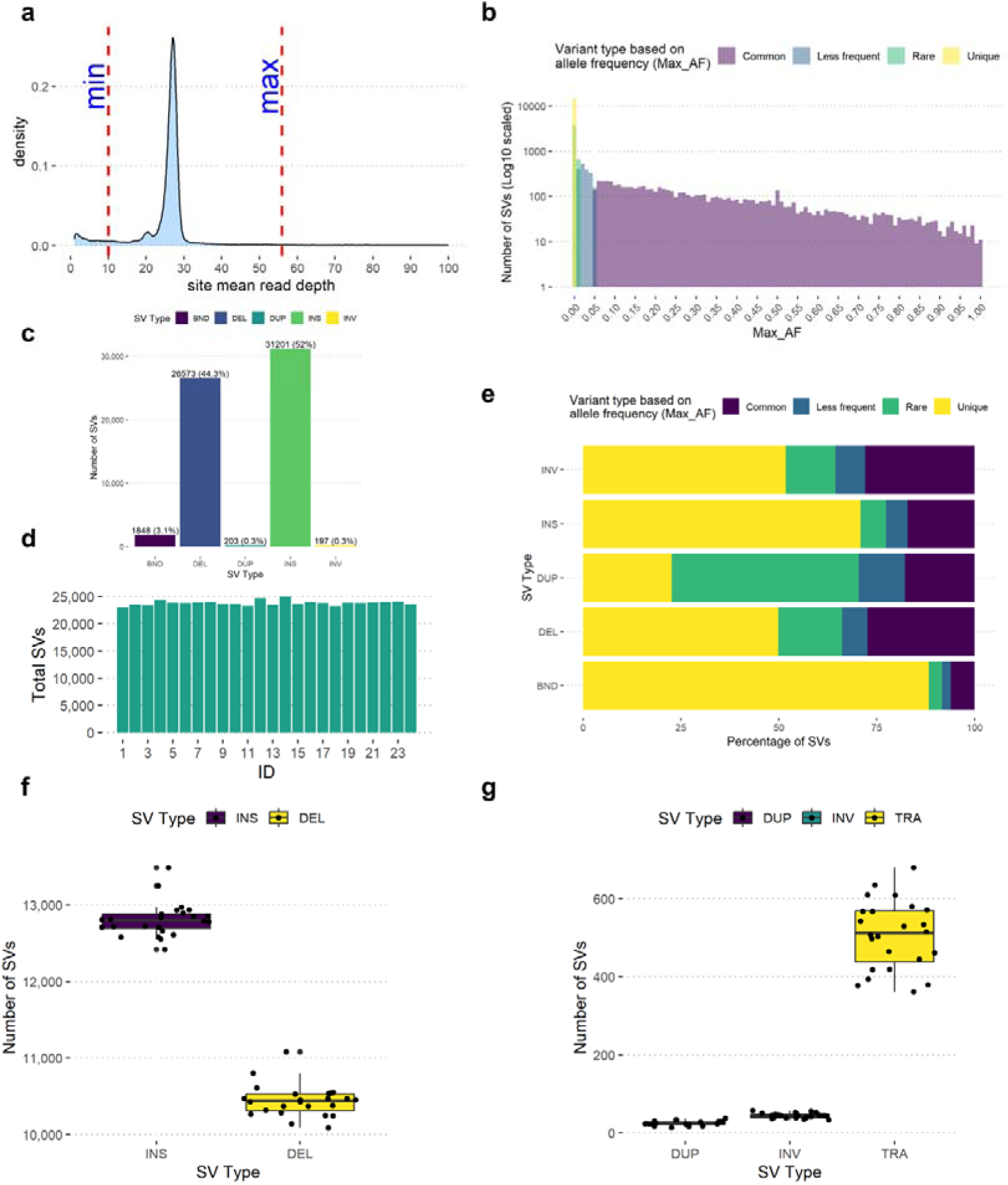
(a) Density plot showing the minimum (10x) and maximum (34x) site mean read depths calculated from a multisample (n=74) VCF file. The minimum (min) and maximum (max) limits are shown via red dotted lines. SV sites below the minimum value and above the maximum value were filtered out. (b) Histogram showing the number of filtered SVs that are common (Max_AF or maximum allele frequency <= 0.05), less frequent (Max_AF < 0.05 and >= 0.01), rare (Max_AF < 0.01 and Max_AF not equal to 0) and unique to the SGP LRS cohort (Max_AF = 0). (c) Bar plot showing the number of filtered SVs stratified by SV type. (d) Bar plot showing the total number of filtered SVs detected per family. (e) Bar plot showing the percentage of common, less frequent and rare SVs based on Max_AF stratified by SV type. (f) Box and jitter plot showing the distribution of the number of INSs (insertions) and DELs (deletions) per family (n=24). (g) Box and jitter plots showing the distribution of the number of filtered DUPs (duplications), INVs (inversions) and TRA (translocations) per family (n=24).

### SV prioritisation and analytical strategy

The aim of the SV prioritisation pipeline was to reduce the resulting data to a manageable number of potentially pathogenic SVs that can be analysed and interpreted given the present understanding and known biology of protein-coding genes. This restricted our analysis to panels of genes that were defined by referring clinicians based on patient phenotype, HPO terms and Genomics England PanelApp diagnostic gene panels [59]. All the parents of the probands in our cohort were unaffected. Based on this understanding, an SV prioritisation strategy was developed to prioritise autosomal *de novo* SVs and SV-affect genes with autosomal recessive MOIs for the condition in question (both homozygous and compound heterozygous), followed by X-linked analysis for affected male patients. In our analysis, whenever an annotated gene overlapped with an SV and the phenotype of the gene matched the patient’s phenotype, the SV was visually inspected and confirmed via Integrative Genomics Viewer [71] (IGV) plots in a systematic manner. This helped rule out false-positive or artefactual SVs incorrectly genotyped by the SV calling and genotyping algorithm because of the misalignment of reads in repeat regions.

We detected a total of 23,024-25,009 SVs genome-wide across all 24 families. A total of 392 SVs genome-wide and 8 within a disease-gene panel were prioritised across different modes of inheritance—autosomal dominant or *de novo* (genome-wide: 229; panel: 4), homozygous autosomal recessive (genome-wide: 45; panel: 0), compound heterozygous (genome-wide: 70; panel: 0), and X-linked (genome-wide: 48; panel: 4), with counts varying across families (Table 4 and Figure 2d). Regardless of whether the proband had an SV, all SVs, which included both rare and common SVs, were included while the SVs were counted after family-based SV calling. The MOI-based filtering ensured that the proband had a rare SV and was primarily responsible for the dramatic reduction in the total number of SVs. We focused on those SVs that had a high likelihood of diagnostic significance in these families and therefore focused our analyses on i) *de novo* SVs in panel genes, ii) SVs in panel genes that could be pathogenic with a biallelic MOI when considered alongside SNVs already detected in SRS data from these families, and iii) SVs that could have an X-linked MOI. We detected four *de novo* SVs in the panel genes (Table 4). Of these, one SV was not considered a true SV after inspection of IGV plots (Family_16, Table 4) and is therefore not discussed further. However, the *de novo* SVs called in families Family_1, Family_3 and Family_11 appeared correct calls on inspection of IGV plots and are described further below as exemplar cases that explain or are most likely to explain the phenotype observed in the proband in these families.

**Table 4:** Summary of high-quality SVs per family by mode of inheritance, genome-wide and within a disease-gene panel. Table 4: Summary of high-quality SVs detected per family, genome-wide and within a disease-gene panel, stratified by mode of inheritance - autosomal dominant/de novo (229; panel: 4), homozygous recessive (45; panel: 0), compound heterozygous (70; panel: 0), and X-linked (48; panel: 4) - with counts varying between families.

| S.No. | Family_ID | Total_SVs | *De Novo* |  | X-linked |  | Autosomal Recessive (Homozygous) |  | Compound Heterozygous |  |
| --- | --- | --- | --- | --- | --- | --- | --- | --- | --- | --- |
|  |  |  | Genome-Wide | Panel genes | Genome-Wide | Panel genes | Genome-Wide | Panel genes | Genome-Wide | Panel genes |
| 1 | Family_1 | 23024 | 2 | 1 | 6 | 0 | 4 | 0 | 1 | 0 |
| 2 | Family_2 | 23527 | 8 | 0 | 0 | 0 | 4 | 0 | 0 | 0 |
| 3 | Family_3 | 23403 | 3 | 1 | 4 | 0 | 0 | 0 | 9 | 0 |
| 4 | Family_4 | 24276 | 2 | 0 | 0 | 0 | 0 | 0 | 0 | 0 |
| 5 | Family_5 | 23869 | 10 | 0 | 1 | 0 | 1 | 0 | 0 | 0 |
| 6 | Family_6 | 23809 | 7 | 0 | 2 | 0 | 2 | 0 | 4 | 0 |
| 7 | Family_7 | 23930 | 5 | 0 | 2 | 0 | 1 | 0 | 1 | 0 |
| 8 | Family_8 | 24001 | 1 | 0 | 4 | 1 | 2 | 0 | 0 | 0 |
| 9 | Family_9 | 23655 | 3 | 0 | 0 | 0 | 1 | 0 | 3 | 0 |
| 10 | Family_10 | 23633 | 2 | 0 | 0 | 0 | 3 | 0 | 2 | 0 |
| 11 | Family_11 | 23374 | 5 | 1 | 0 | 0 | 0 | 0 | 2 | 0 |
| 12 | Family_12 | 24656 | 13 | 0 | 0 | 0 | 0 | 0 | 5 | 0 |
| 13 | Family_13 | 23499 | 4 | 0 | 0 | 0 | 5 | 0 | 0 | 0 |
| 14 | Family_14 | 25009 | 7 | 0 | 5 | 0 | 0 | 0 | 29 | 0 |
| 15 | Family_15 | 23650 | 4 | 0 | 0 | 0 | 0 | 0 | 0 | 0 |
| 16 | Family_16 | 24012 | 36 | 1 | 0 | 0 | 3 | 0 | 1 | 0 |
| 17 | Family_17 | 23797 | 3 | 0 | 5 | 1 | 5 | 0 | 0 | 0 |
| 18 | Family_18 | 23273 | 2 | 0 | 5 | 2 | 4 | 0 | 7 | 0 |
| 19 | Family_19 | 23863 | 9 | 0 | 0 | 0 | 0 | 0 | 1 | 0 |
| 20 | Family_20 | 23819 | 14 | 0 | 0 | 0 | 1 | 0 | 1 | 0 |
| 21 | Family_21 | 23917 | 17 | 0 | 4 | 0 | 2 | 0 | 1 | 0 |
| 22 | Family_22 | 24001 | 14 | 0 | 5 | 0 | 1 | 0 | 1 | 0 |
| 23 | Family_23 | 24041 | 46 | 0 | 0 | 0 | 3 | 0 | 1 | 0 |
| 24 | Family_24 | 23609 | 12 | 0 | 5 | 0 | 3 | 0 | 1 | 0 |

Genome-wide *de novo* SVs were additionally detected, and the number of *de novo* SVs detected was greater than expected. Each generation is expected to have at most one true *de novo* SV [21]. However, the bioinformatics pipeline detected up to 46 SVs in the families under study. A manual examination of these *de novo* SV calls was needed to rule out false positives and, if necessary, validate them via additional orthogonal approaches. Additional File 1: Supplementary Figure 6 shows a scatter plot of the number of genome-wide *de novo* SVs and the mean family depth. Two main clusters are clearly formed where families sequenced at a greater depth (R9.4.1+R10.4.1) have a lower number of genome-wide SVs than the second cluster, which contains families sequenced at a lower depth (only R9.4.1). Sequencing at a greater depth therefore increases the chances of sampling reads of the alternate allele, which might be missed at a lower depth. Furthermore, establishing the clinical relevance of the affected gene could rely on new gene discovery for a particular phenotype, which is beyond the scope of this study.

The statistics on total SVs per family stratified by different SV types are shown in Additional File 1: Supplementary Table 7, Figure 2f and g, with no clear outliers.

Three families contained X-linked recessive SVs in their panel genes. Family_8 (recruited disease: intellectual disability and syndromic congenital heart disease) contained a maternally inherited insertion in *CDKL5*; however, this was not pursued further, as the gene has an X-linked dominant MOI in OMIM for epileptic encephalopathy early infantile type 2, and the mother of the proband is unaffected. Family_17 also had an *Alu* insertion in the noncanonical transcript of *PHEX, a* gene linked to X-linked dominant hypophosphatemic rickets. As this does not match the patient’s phenotype (recruited disease: Untra-rare undescribed monogenic disorders) and the mother is unaffected, this insertion was not pursued further. Family_18 had two X-linked insertions, one in the intronic region of *DMD* (330 bp *Alu* INS in an antisense manner) and another downstream of *BCOR* (82 bp INS, not in the promoter). None of the phenotypes associated with these genes matched the affected proband’s phenotypes (recruited disease: syndromic congenital heart disease and syndromic cleft lip and/or cleft palate). Additional examination of chromosome-wide X-linked SVs did not suggest any further potential diagnoses, as most of the SVs were either intergenic or intronic, or the gene phenotype did not match the patient’s phenotype. Establishing the clinical relevance of these SVs is beyond the scope of this report, which would require additional functional studies with either RNA-Seq or methylation analysis that would need to be the work of future studies.

None of the families had any SVs overlapping a biallelic panel gene with autosomal homozygous recessive MOI (Table 4). Additional examination of genome-wide SVs at this MOI revealed that they were either intergenic or intronic or that the gene’s phenotype was not a good match with the patient’s phenotype. No compound heterozygous scenarios (SV+SNVs) were found in any of the biallelic panel genes associated with a given family.

A compilation of genome-wide *de novo* SVs and compound heterozygous instances (SV+SNVs) is shown in Additional File 2: Supplementary Tables 8 and 9, respectively, and is currently being investigated for novel causal gene identification related to rare disease phenotypes.

### Exemplar cases

Three examples of this SGP LRS study are described below, in which pathogenic or likely pathogenic SVs were identified. IGV plots made via LRS data showing the SV findings in the three families are shown in Additional File 1: Supplementary Figure 7. The molecular changes in exemplar families 1 and 2, when analysed alongside the proband phenotype, were accepted as pathogenic at multidisciplinary team meetings (MDTs) of clinical geneticists and clinical scientists. The changes identified in exemplar Family 3 were designated as likely pathogenic.

### Exemplar Family 1 (Recruited Disease: Intellectual disability)

The proband is the firstborn child of non-consanguineous parents, born at term with a birth weight of 3.5 kg (−0.67 s.d.). In early childhood, there were concerns about poor feeding and delayed developmental milestones. The proband sat unsupported in infancy and achieved independent walking within the first 5 years of life. The parents and younger siblings had no health issues.

The proband was referred to Clinical Genetics at the age of 5-9 years for an undiagnosed neurodevelopmental disorder and recent onset of seizures. Reported medical issues included unclassified epilepsy, developmental delay (nonverbal), microcephaly (−3.5 s.d.), sleep onset and persistence disorders, skeletal abnormalities (finger contractures, bilateral calcaneal valgus deformity causing chronic foot pain), and convergent squint. On examination, the proband was found to be microcephalic with a flattened occiput. Facial features included convergent squint, mid-face hypoplasia, micrognathia, a prominent nose with flattened nasal bridge, a short philtrum, prominent ears, and thickened lips. Striking finger contractures and a contracted right elbow were also noted.

Clinical investigations included normal magnetic resonance imaging (MRI) of the brain (ages 5-9 years), no specific abnormalities detected on electroencephalogram (EEG) (ages 5-9 years) and no abnormalities found on a basic metabolic screen (thyroid function test, creatine kinase, lead, urate, chitotriosidase and biotinidase levels). Previous genetic investigations did not reveal a cause and included a chromosome microarray as well as recruitment for the Deciphering Developmental Disorders (DDD) study, through which exome sequencing was performed without a diagnosis.

The proband was subsequently recruited to SGP, where initial analysis via SRS did not identify a causative variant in the analysis of Tier 1 (predicted protein-truncating) or Tier 2 (predicted protein-altering) variants.

SGP LRS detected one event that was a 1.4 Mb *de novo* inversion that overlapped with a candidate gene for numerous neurodevelopmental and neurological disorders designated *AUTS2* or autism susceptibility candidate 2 (MIM: 615834) encoding Activator of Transcription and Developmental Regulator protein [72]. It particularly helps in neurogenesis and maturation of neurons and plays a key role in synapse formation and dendritic spine regulation [73, 74]. One of the inversion breakpoints was located within intron 2 of the gene and was predicted to have a high functional impact on the protein-coding sequence, resulting in the removal of exons 1 and 2. This disruption likely impairs gene expression and provides a molecular explanation for the proband’s phenotype described above. The same *de novo* inversion was also prioritised via SVRare analysis of SRS data [75], confirming the SV via a second orthogonal approach. A samplot [76] image of the *AUTS2* inversion is shown in Figure 3, and an IGV plot is presented in Additional File 1: Supplementary Figure 7a.

**Figure 3:**
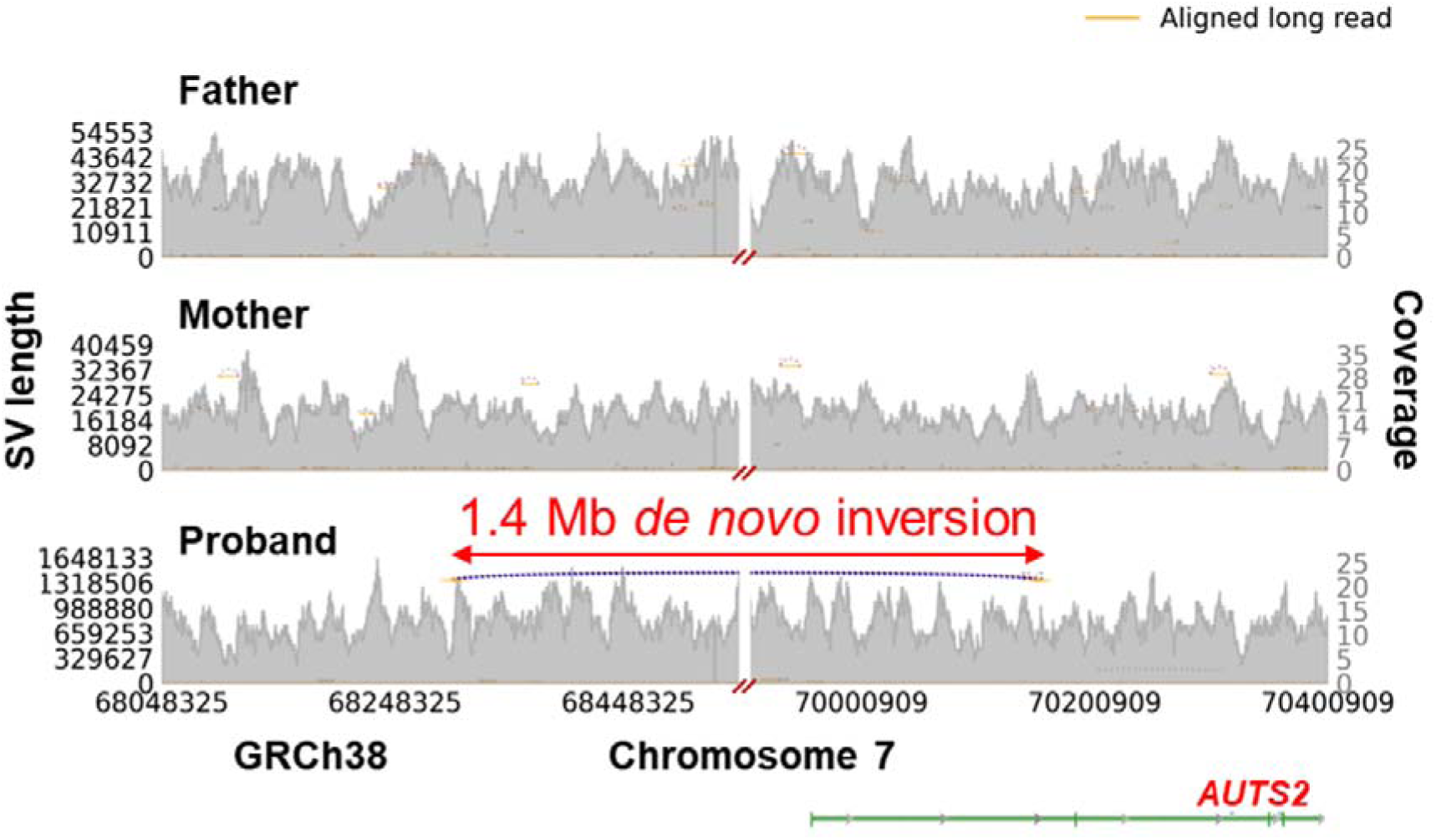
A 1.4 Mb *de novo* inversion found from LRS data in the proband from exemplar family 1 that overlaps with *AUTS2* and is absent in the parents. The inversion is denoted by a purple dotted line in the proband. Other smaller SVs can be ignored. The x-axis refers to the chromosome base position. The left y-axis refers to the SV length, and the right y-axis represents the depth of coverage.

### Exemplar Family 2 (Recruited Disease: Ultrarare undescribed monogenic disorders)

The proband with unaffected parents had three-limb ectrodactyly affecting both hands and the right foot and postaxial polydactyly of the left foot. Retrognathia and overfolded ear helices were also noted. There was dental overcrowding, which was treated with tooth extractions and a maxillary osteotomy. Audiometry in adulthood revealed narrow ear canals and mild bilateral conductive hearing loss. Resting sinus bradycardia was noted, but the heart was structurally normal.

Ectrodactyly was also observed antenatally in the proband’s child. The child was born prematurely at 32-36 weeks. A patent ductus arteriosus, an atrial septal defect (ASD) and a thickened pulmonary valve were noted at birth. The ductus arteriosus and ASD subsequently closed spontaneously, but pulmonary valve stenosis persisted with a degree of pulmonary regurgitation and sinus bradycardia. The pinnae were small but normally formed, with normal ear canals. Audiometry revealed fluctuating mild to moderate conductive hearing loss, which did not require treatment. Additional features included mild retrognathia with dental overcrowding, delayed closure of the anterior fontanelle, and several fractures with minor trauma. A DEXA bone scan between ages 10-14 years demonstrated a reduced bone density Z score of -4.0, although it was unclear if this was related to the other clinical features.

For this family, LRS was carried out for the trio of the first and second generations (unaffected parents and the proband). SRS was performed for the trio and for the affected proband’s affected child. Using trio LRS and SV analysis, a *de novo* inversion was discovered in the split hand/foot malformation type 1 (*SHFM1*, MIM: 183600) region, in which several pathogenic SVs that are causative of limb malformation have been reported in the literature (Figure 4b) [77–80]. This event was prioritised incidentally because one of the panel genes (*COL1A2*) was present in the middle of the inversion. However, since none of the inversion breakpoints were located within the panel gene, it would not have an impact on the gene’s expression. When genes nearest to the breakpoints were examined, two genes, *DLX5* and *DLX6*, which are important genes in the WNT pathway and play critical roles in limb development [79], were observed to be present very close to the inversion’s distal breakpoint. Cytoplasmic dynein intermediate chain 1 (*DYNC1I1)* is located at one end of the inversion and contains enhancer elements regulating the expression of *DLX5* and *6*. It is hypothesised that the inversion displaced the regulatory elements far from the DLX5/6 genes, leading to their downregulation, affecting the limb development process and causing split hands in the patient [77–80]. On extending the analysis to the SRS data of the proband’s child, who also has ectrodactyly, it was observed that the child inherited the inversion from the affected proband (Figure 4a,c). This event was also detected retrospectively in the SRS data using SVRare providing orthogonal confirmation. The IGV plot of the LRS data of this family (only the proband and parents) is presented in Additional File 1: Supplementary Figure 7b.

**Figure 4:**
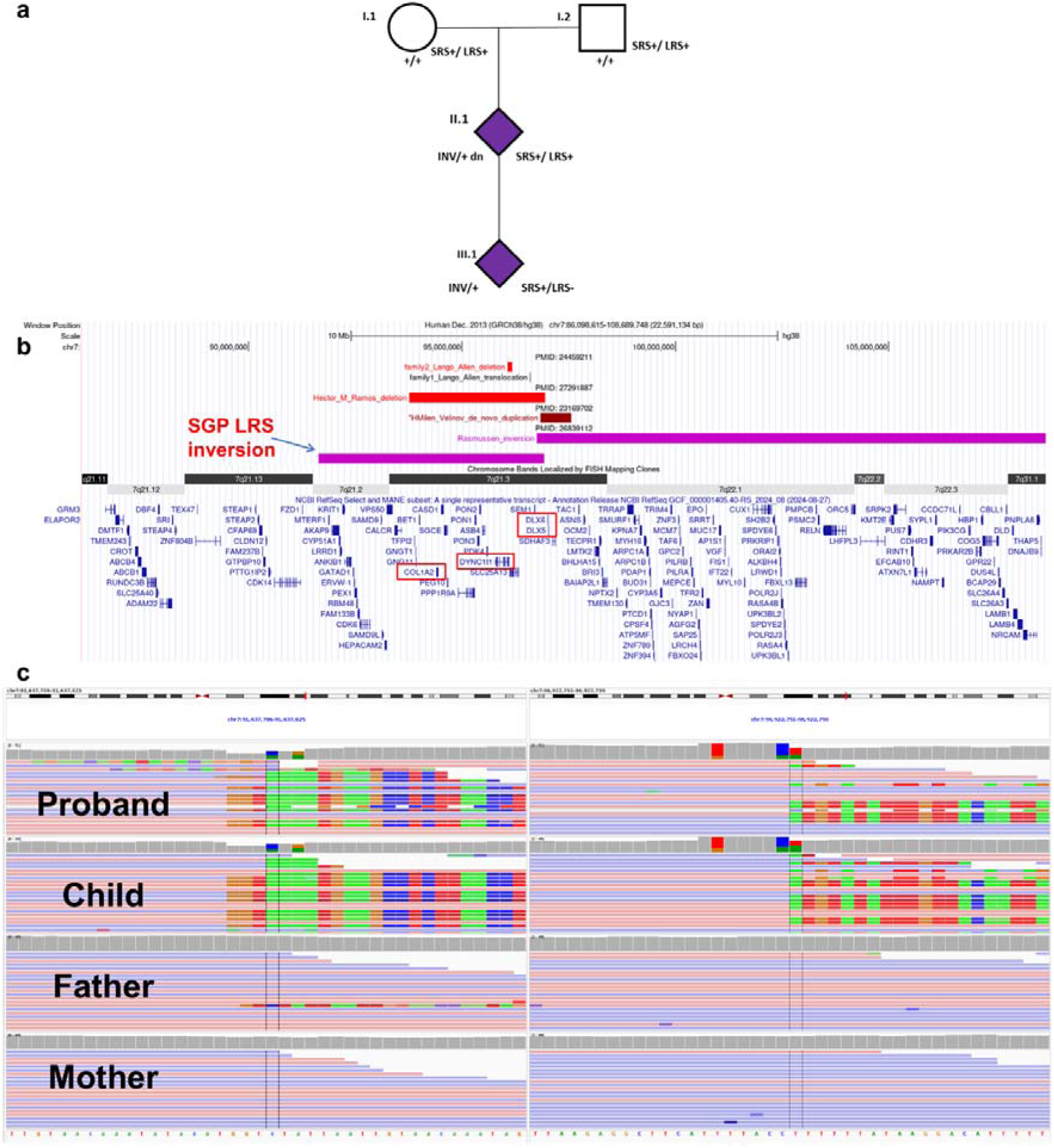
(a) Pedigree diagram showing the three generations of exemplar family 2. Circle and square symbols represent unaffected females and males, respectively, while a diamond indicates masked or undisclosed sex. SRS and LRS labels along with the symbols “+” or “-” indicate the availability and unavailability of the type of sequencing data, respectively. The proband (II.1) has a heterozygous *de novo* inversion (denoted by “dn”) near the DLX5/6 locus, which was inherited by the child (III.1). Affected individuals with DLX5/6 inversion are shaded in purple. (b) The UCSC session displays the SHFM1 (split hand-foot mouth) locus in the 7q21 region. The coloured bars show various SVs (red represents deletions, brown represents duplications, and purple represents inversions) that have been reported previously, along with the inversion present in the patient from the SGP LRS cohort. The track names denote the type of SV along with the names of the authors of the publications they are described in, whereas the description on the top of the SVs shows the PubMed ID number of the publication. The red boxes indicate the genes of interest described in the main text. (c) IGV plot showing soft-clipped bases near the inversion breakpoint in the proband (II.1) and proband’s child (III.1), which are absent in the parents. The plot was generated from SRS data in the Genomics England (GEL) research environment, which also included sequencing of the child (III.1).

### Exemplar Family 3 (Recruited Disease: Unexplained skeletal dysplasia)

The proband is the only child of nonconsanguineous healthy parents and was born at term by forceps delivery. Family history and early development were unremarkable. The proband presented in childhood (ages 5-9 years) with an abnormal gait and bilateral hip pain that developed in the left and right hips between 5-9 years. Other joints were initially unaffected, and a diagnosis of bilateral Perthes disease was suspected. Progressive contractures, which increasingly restricted mobility, subsequently affected the hips from early adolescence (ages 10-14 years) and the elbows from mid-adolescence (ages 15-19 years). From mid-adolescence (ages 15-19 years), there was progressive pain and swelling of the interphalangeal finger joints, for which the proband received intra-articular injections. There was no history of bruising tendencies, joint laxity or joint dislocations.

On examination, there was a reduced range of hip joint movement, significant weakness of his pelvic girdle musculature but normal muscle power elsewhere, with normal reflexes in his upper and lower limbs. The proband walked with thoracic kyphosis, lumbar lordosis, flexed knees and stiff hips. Height (146 cm in early adolescence, ages 10-14 years), proportion, vision, hearing and teeth were normal, and the proband was non-dysmorphic. Clinical investigations, following orthopaedic review, included radiology with skeletal survey X-rays (ages 10-14 years) showing a slightly ovoid shape of some vertebral bodies. MRI of the pelvis revealed atrophy of the pelvic musculature but no evidence of myositis or myopathy. MRI of the hips revealed fragmented, collapsed, irregular proximal femoral epiphyses. A subsequent review of his skeletal survey revealed bilateral symmetrical irregular sclerosis of the femoral epiphyses, mild metaphyseal broadening with minor cystic changes and subchondral fracturing and collapse, in addition to bilateral hip joint effusions. Some end-plate concavities of the lumbar spine were also noted. The bone age was appropriate. A diagnosis of multiple epiphyseal dysplasia (MED) was considered.

Sanger DNA sequencing of 5 MED-related genes (*COMP, COL9A1, COL9A2, COL9A3 and MATN3*), in addition to *LMNA,* was performed, but no pathogenic variant was identified. Muscle biopsy and pathology (including ultrastructural studies) revealed nonspecific findings, with only atrophic fibres and no evidence of a mitochondrial respiratory chain defect. Serum creatine kinase (CK) levels, nerve conduction studies, electromyograms and lumbar punctures were all normal. Chromosomal microarray revealed a *de novo* sub microscopic heterozygous deletion of approximately 55 kb at *2q35*, resulting in a multiexon deletion of the FN1 gene (MIM: 184255), which was considered a variant of uncertain significance (VUS). Additionally, the SGP SRS study utilised gene panels covering skeletal dysplasia, limb girdle muscular dystrophy, distal myopathies, arthrogryposis, congenital myopathy and congenital muscular dystrophy. No causative variant was identified on initial sequencing analysis.

From the LRS in this study, a *de novo* deletion event was detected in the proband, a multiexon in-frame deletion spanning exons 6 to 41 (out of a total of 46 exons in the MANE transcript) in *FN1* (Fibronectin 1) of 59.8 kb in length (Figure 5, Additional File 1: Supplementary Figure 7c), reconfirming the chromosomal microarray experiment and providing the exact and precise breakpoints of the deletion. The deletion was visually inspected in IGV, where a clear drop in the depth of coverage was detected in the proband with respect to the father’s and mother’s depth of coverage in the same region (Figure 5). The deletion was also prioritised after SVRare analysis of the SRS data. *FN1* is an important gene for the musculoskeletal system and encodes a multimeric glycoprotein in the extracellular matrix that is involved in cell adhesion and migration [81]. Importantly, the *FN1* deletion was initially genotyped as homozygous by LRS, although visual inspection in IGV/samplot confirmed that it was heterozygous. This discrepancy led to it being filtered out when a stringent allele frequency (AF) threshold of 0.01 (see Methods) was applied to identify *de novo* variants. However, when a slightly more lenient threshold of 0.015 was used, the variant was retained and shortlisted. This highlights both a genotyping error by the variant caller and the sensitivity of variant prioritisation to AF thresholds. The *FN1* SV was reclassified as likely pathogenic, and additional work is ongoing in establishing a definitive connection between *FN1* and bilateral Perthes disease.

**Figure 5:**
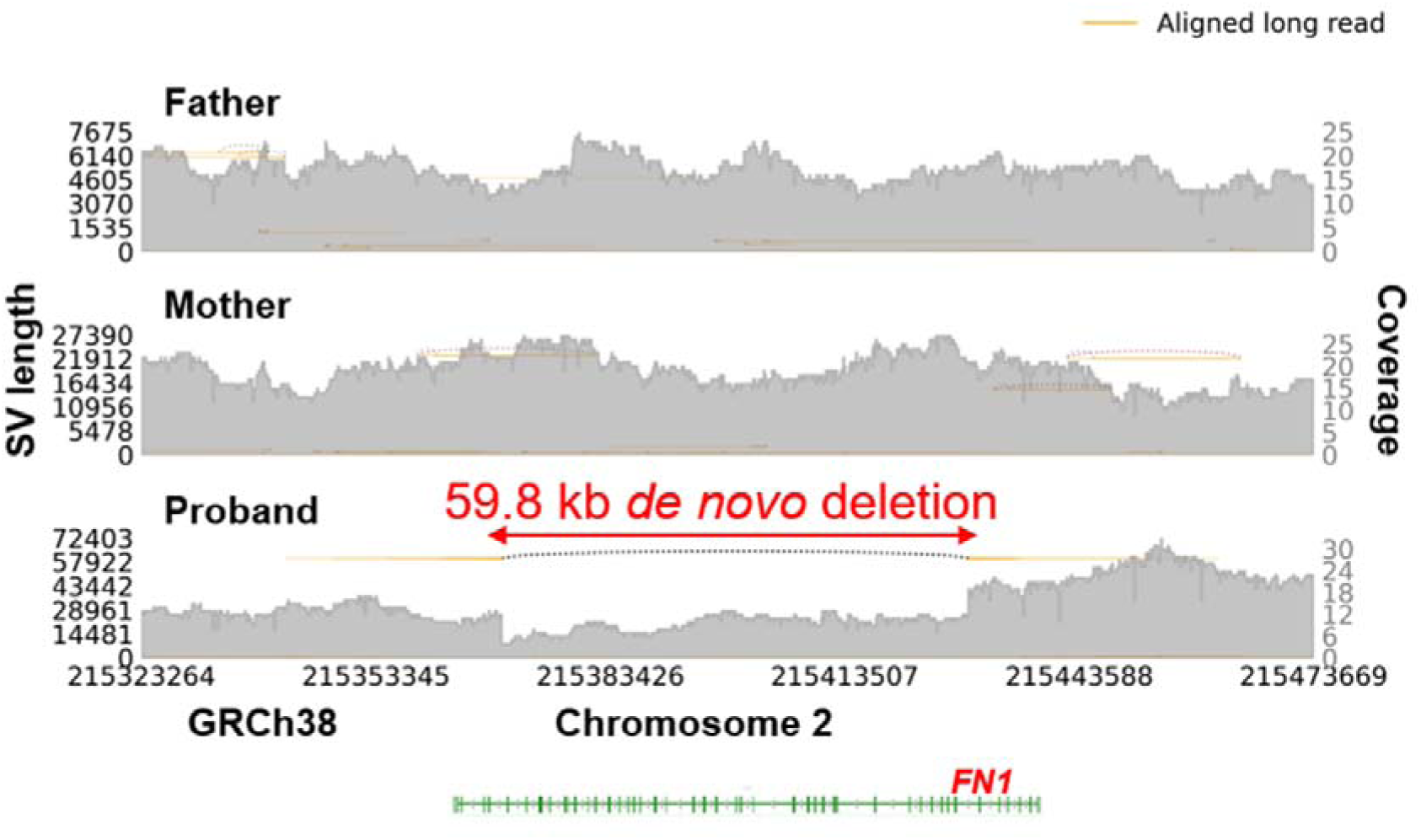
A 59.8 kb *de novo* deletion found from LRS data in the proband from exemplar family 3, which is absent in both parents. The multiexon deletion is found within *FN1*, with exons 6 to 41 of the MANE transcript deleted. The deletion is denoted by a black dotted line in the proband. The heterozygous deletion can be visualised by the lower depth of coverage between the breakpoints of the SV. Other smaller SVs can be ignored. The x-axis refers to the chromosome base position. The left y-axis refers to the SV length, and the right y-axis represents the depth of coverage.

## Discussion

In this study, we benchmarked presently available software for the analysis of long-read sequences to detect structural variation, defined requirements for sample processing and DNA integrity, and applied optimal software combinations for SV analysis of panel genes in a cohort of 74 individuals from 24 families. These analyses of LRS, coupled with an additional analysis in these families to prioritise SVs from SRS data as an orthogonal approach, enabled the diagnosis or likely diagnosis in 3 of the 24 families in this cohort. Additional genome-wide SV analysis, with the aim of identifying further pathogenic SVs, is currently ongoing via a panel-agnostic approach.

The analytical tools for SV detection, both for LRSs and SRSs, are developing rapidly. Multiple benchmarking studies examining various aligner and SV caller combinations have been published previously [39, 40]. For SV detection in LRS, we benchmarked the variant calling process via three long-read-based aligners (minimap2, NGMLR and lra) and two dedicated long-read SV callers (cuteSV and Sniffles2). The two SV callers were specifically chosen because, in addition to single-sample SV calling, they also allow either force calling or multisample methods while calling SVs (or both), and in this study, the clinical samples are trios/quads. For single-sample SV calling, a high F1 score led to the selection of the minimap2 (for alignment) and cuteSV (for SV calling) combination for the initial pilot study in terms of SV and its genotype calling. For family-based SV calling, Sniffles2 was chosen to call SVs based on Mendelian inconsistency and the degree of missingness.

To assess the importance of DNA quality and fragment size, our pilot study examined the impact of size selection and the DNA quality of the input DNA samples. With size selection, partially degraded DNA gave comparable read coverage and SV calls to DNA samples of greater integrity. Similarly, size selection, while increasing the proportion of reads that were greater than 10 kb, did not significantly alter the SV calling rate. Notably, current ONT protocols that utilise R10.4.1 flow cells do not require shearing of DNA, as was used in this study. Nevertheless, a major difference was observed in sequence read depth, with the application of a single sample per flow cell, between the R9.4.1 and R10.4.1 flow cells and with improved chemistry (Kit V9 and Kit V14, respectively), with increased depth of coverage and consequently improved accuracy of SV calling/genotyping at deeper depths of coverage.

A key determinant of accurate identification of pathogenic SVs is the ability to comprehensively detect all genome-wide SVs, reliably distinguish true positives from false positive SV calls and effectively filter out high-frequency SVs common to the general or study population. We undertook several automated and manual approaches to filtering SV calls, including in this study, the primary restriction of variant analysis within relevant gene panels (although all genome-wide SVs were generated), manual inspection of IGV plots to remove false positive calls, and interrogation of various public databases, including SV population frequency databases, ClinVar and OMIM, to determine likely pathogenic variants according to the documented phenotype of individual patients and family members. In addition to the above processes, we searched for compound heterozygotes where SVs in the LRS and SNVs in the SRS might indicate pathogenicity in genes with an autosomal recessive mode of inheritance. We also screened the whole X chromosome for SVs, which might explain the rare disease phenotype in male cases where the SV was also present in the proband’s mother. Finally, to extend the previous analyses in the SRS, SVRare software was applied to prioritise SVs in the SRS data. Ultimately, from approximately 23,000 to 25,000 called SVs per family, we were able to filter down to 1-46 SVs per family when only those SVs were retained according to the MOI, as described in the Methods.

Several steps were taken in the SV calling and filtering processes to maximise calling accuracy, to remove common SVs that were not pathogenic and to optimise the likelihood of detecting clinically relevant pathogenic SVs. These included increasing the read coverage by additional sequencing on the R10.4.1 platform, adjusting the population and cohort-specific allele frequencies to maximise true SV detection, validation of SVs detected in the LRS by specifically seeking to confirm the presence of these SVs in the previously generated SRS, and individual comparison of documented gene-specific phenotypes with the patient-specific phenotypes in our families. To maximise the opportunity for detecting pathogenic SVs in this cohort, in which the parents of all probands were unaffected, we chose to focus primarily on *de novo* SVs within panel genes and panel gene regions, on the combination of SVs and SNVs in panel genes with an autosomal recessive MOI, on inherited X-linked genes in male probands where the mother was unaffected but carried the X-linked SV, and on homozygous SVs in genes with autosomal recessive MOIs.

These analytical decisions enabled the new finding of two pathogenic variants, one at the *DLX5/6* locus and one in *AUTS2*. The description of a novel *DLX5/6* inversion linked to ectrodactyly helps to build on the literature that defines the critical regulatory regions required for limb development [77–80]. The involvement of *AUTS2* in a range of neurodevelopmental disorders, dysmorphic features, and skeletal abnormalities is well established, supporting its role in explaining the proband’s observed phenotype [74, 82, 83]. Additionally, one likely pathogenic variant was identified in *FN1*. The known *FN1* gene phenotype, coupled with the respective phenotype in the proband, showed a close alignment between the described gene and patient phenotypes, with the exception that even after expert radiological review of the proband’s X-rays, the patient’s phenotype did not include corner fractures, which are usually considered an invariant feature of *FN1* mutation. This led to the conclusion after multidisciplinary team appraisal that the *FN1* SV in this patient should be categorised as likely pathogenic. Further clinical investigations and discussions are ongoing to establish the role of *FN1* in its involvement in causing bilateral Perthes features, which was the patient’s original diagnosis.

Our focus on panel genes, while maximising the possibility of identifying pathogenic variants at this stage of our analyses, increases the likelihood of the discovery of further pathogenic variants, for example, by examining SVs in the remainder of the genome, which may identify pathogenic SVs in newly discovered rare disease genes. In addition, the focus of this study was primarily on coding sequence variation, which raises the possibility that SVs in upstream or downstream noncoding regions of panel genes, or within genes elsewhere in the genome, may also identify further pathogenic variants in this cohort. Additional sequencing modalities also help enhance the diagnostic yield when variants in question are present in the noncoding region of the genome. We are currently carrying out further analyses of the LRS, including investigations of the SVs identified in the rest of the genome, identification of DNA methylation abnormalities and, where samples are available, transcript analysis through RNA-seq of leukocyte RNA. Exemplar cases have been described in a recent study where analysis of DNA methylation, available directly from the ONT sequence, was able to prioritise noncoding SVs as orthogonal validation for the pathogenicity of a deep intronic SNV [84]. More extensive LRS multiomics profiling studies have also helped resolve the molecular basis of a patient’s disease [85].

We recognise that our study has limitations. First, not all the samples were sequenced on the R10.4.1 platform, which has a higher yield and performance than the R9.4.1 platform. Therefore, updating all samples to a coverage of 30--40x on the R10.4.1 platform could lead to new diagnoses because of sampling bias with an increased likelihood of false genotype calls at lower sequence coverage. Second, although we primarily searched for panel genes for SVs, further analysis of genes outside the panel could lead to additional new diagnoses. Although we do have genome-wide SV data stratified by different MOIs, additional in-depth examination of the SVs is further needed. For example, by prioritising genome-wide *de novo* SVs, we could look for any SV overlapping a haploinsufficient gene. This might lead to the discovery of new causal genes underlying rare disease phenotypes, which would require additional functional validation. To aid this, we catalogued the genome-wide *de novo* SVs and compound heterozygous (SV+SNV) events identified in the families in this study (Additional File 2: Supplementary Tables 8 and 9, respectively). We explored genome-wide and chromosome-wide SVs for homozygous autosomal recessive and X-linked MOIs, respectively, but no SVs that could explain the affected proband’s phenotype were identified. Furthermore, although the most recent versions of the gene panels were used for this study at the time of analysis, updating to the very newest versions could add genes that have recently been linked to human disease because PanelApp diagnostic gene panels are updated frequently [59].

Finally, with the rapid advancement of available software used in our analyses, it is possible that updates to the software or the use of new machine learning-based algorithms for SV discovery (such as SVcnn [86]) could identify additional pathogenic variants in this cohort. However, notably, not all available software offers multisample/family-based SV calling methodology, which was essential in our analysis.

The exemplar cases described in this cohort complement but do not necessarily outperform SRS, as we focused on this study, mainly on panel genes. Future studies should be hypothesis-free (panel-agnostic) and fully genome-wide. LRS broadens the search space for SVs by detecting more variants than SRS. [55, 87, 88]. This broader detection enables the exploration of additional SVs that may explain a patient’s phenotype, particularly when no pathogenic SVs are identified in panel genes.

## Conclusions

There have been multiple studies where reanalysis of genomes via LRS has facilitated additional diagnosis in previously unsolved cases [31, 33, 36, 89, 90] because they can seamlessly map in regions where SRS struggles. We therefore conclude that the ability to comprehensively identify SVs across the genome in both individual- and family-based samples through long-read sequencing on platforms such as ONT, in tandem with increasingly sophisticated software for identifying such structural variation, creates new opportunities for making molecular genetic diagnoses in cases and families with rare diseases. In this study, we identified two pathogenic structural variants and one likely pathogenic structural variant across three families, bringing the diagnostic odyssey to a close for at least two of them (*AUTS2* and *DLX5/6*) out of a total of 24 families. Further analyses with the existing LRS and SRS remain possible, but these exemplars indicate how additional diagnoses can be made in rare disease families with the approaches used in this study.

**Abbreviations**
SRS: Short-read sequencing
WGS: Whole-genome sequencing
SGP: Scottish Genomes Partnership
GEL: Genomics England
LRS: Long-read sequencing
SVs: Structural variants
WES: Whole-exome sequencing
DIN: DNA integrity number
HMW: High-molecular-weight
ONT: Oxford Nanopore Technologies
TP: True-positive
FP: False-positive
VCF: Variant Call Format
GT: Genotype
QUAL: Phred-scaled quality score
GQ: Phred-scaled genotype quality
BAM: Binary Alignment/Map
LINE-1: Long interspersed nuclear elements
SINEs: Short interspersed nuclear elements
QC: Quality control
VEP: Variant Effect Predictor
AF: Alternate Allele frequency
Max_AF: Maximum Alternate Allele frequency
HPO: Human Phenotype Ontology
INS: Insertions
DEL: Deletions
DUP: Duplications
INV: Inversions
TRA: Translocations
BND: Breakend
IGV: Integrative Genomics Viewer
MOI: Mode of inheritance
OMIM: Online Mendelian Inheritance in Man
CCDG: Center for Common Disease Genomics
1000G: 1000 Genomes
TOPMed: Trans-Omics for Precision Medicine
SNV: Single nucleotide variant
MRI: Magnetic resonance imaging
EEG: Electroencephalogram
DDD: Deciphering Developmental Disorders
ASD: Atrial septal defect
WNT: Wingless-related integration site
DEXA: Dual-energy X-ray Absorptiometry
CK: Serum creatine kinase
VUS: Variant of uncertain significance
MANE: Matched Annotation from NCBI and EMBL-EBI
UCSC: University of California, Santa Cruz

## Declarations

### Ethics approval and consent to participate

Samples were collected from 73 individuals for the long-read sequence study within the Scottish Genomes Partnership programme project “NHS Scotland in 100,000 genomes study”. All gave informed consent for use of samples and phenotype data for research purposes, including genome sequencing, data store and de-identified data sharing. This research study was approved by North of Scotland Research Ethics Committee (16/NS/0137) and Scotland A Research Ethics Committee (17/SS/0113); the Public Benefit and Privacy Panel (1516-0377); and NHS Scotland health board Research and Development departments.

### Consent for publication

Pedigree details and details of the clinical phenotype were included for three families, all of whom gave additional consent for inclusion of their data and clinical features in this publication.

### Availability of data and materials

Illumina and ONT genome sequencing data related to this study is available in the National Genomic Research Library (NGRL) (https://doi.org/10.6084/m9.figshare.4530893.v7). Details of how to access these data are available at https://www.genomicsengland.co.uk/join-us. Access will be provided via Amazon WorkSpaces through project id “RR566”. For academic researchers, host institutions also need to sign a formal agreement. The codes and custom scripts used in this manuscript will be made available upon request.

**GRCh37 human reference genome and index:**

https://ftp-trace.ncbi.nlm.nih.gov/ReferenceSamples/giab/release/references/GRCh37/hs37d5.fa.gz

https://ftp-trace.ncbi.nlm.nih.gov/ReferenceSamples/giab/release/references/GRCh37/hs37d5.fa.gz.fai

**GRCh38 human reference genome and index:** ftp://ftp.ncbi.nlm.nih.gov/genomes/all/GCA/000/001/405/GCA_000001405.15_GRCh38/seqs_for_alignment_pipelines.ucsc_ids/GCA_000001405.15_GRCh38_no_alt_pl us_hs38d1_analysis_set.fna.gz

ftp://ftp.ncbi.nlm.nih.gov/genomes/all/GCA/000/001/405/GCA_000001405.15_GRCh38/seqs_for_alignment_pipelines.ucsc_ids/GCA_000001405.15_GRCh38_no_alt_pl us_hs38d1_analysis_set.fna.fai

**PanelApp website:** https://panelapp.genomicsengland.co.uk/

## Author contributions

T.J.A. conceived the study, obtained funding and wrote the manuscript; P.D. designed the bioinformatics analysis methodology, performed data analysis on all LRS samples and wrote the manuscript with inputs from T.J.A., J.C.T., A.T.P., C.R., A.M., M.H. and J.S.L.; J.Y. developed SVRare software and J.Y. and A.T.P. performed the analysis of SRS SV data via SVRare; Z.M. oversaw the governance and permissions aspects of the study; A.E.F.M. performed data analysis on SRS and LRS SV data for the compound heterozygous analysis; C.N., M.T., U.T. and J.S.L. undertook the library preparation and sequencing of all clinical samples; A.R., E.S.T. and R.M. provided clinical and phenotypic details for the cases described in the manuscript; Z.M., M.A., D.B., J.B., T.B., V.C., A.D., A.L. and N.W. recruited patients and obtained phenotypic details from study participants.

## Data Availability

Illumina and ONT genome sequencing data related to this study is available in the National Genomic Research Library (NGRL) (https://doi.org/10.6084/m9.figshare.4530893.v7). Details of how to access these data are available at https://www.genomicsengland.co.uk/join-us. Access will be provided via Amazon WorkSpaces through project id 'RR566'. For academic researchers, host institutions also need to sign a formal agreement. The codes and custom scripts used in this manuscript will be made available upon request.
GRCh37 human reference genome and index:
https://ftp-trace.ncbi.nlm.nih.gov/ReferenceSamples/giab/release/references/GRCh37/hs37d5.fa.gz
https://ftp-trace.ncbi.nlm.nih.gov/ReferenceSamples/giab/release/references/GRCh37/hs37d5.fa.gz.fai
GRCh38 human reference genome and index: ftp://ftp.ncbi.nlm.nih.gov/genomes/all/GCA/000/001/405/GCA_000001405.15_GRCh38/seqs_for_alignment_pipelines.ucsc_ids/GCA_000001405.15_GRCh38_no_alt_plus_hs38d1_analysis_set.fna.gz
ftp://ftp.ncbi.nlm.nih.gov/genomes/all/GCA/000/001/405/GCA_000001405.15_GRCh38/seqs_for_alignment_pipelines.ucsc_ids/GCA_000001405.15_GRCh38_no_alt_plus_hs38d1_analysis_set.fna.fai
PanelApp website: https://panelapp.genomicsengland.co.uk/

## Acknowledgements

This study would not have been possible without families, patients, clinicians, nurses, research scientists, laboratory staff, informaticians and the wider Scottish Genomes Partnership team, to whom we extend gratitude. We thank Asier Gonzalez, Olga Leonova and Elliot Gould from Genomics England (GEL) for their inputs and support with data uploaded to the NGRL; Ailin Buzzi from GEL for inputs on data governance and ethics; and Adam Giess (GEL) for early technical discussions on data analysis. We thank Oxford Nanopore Technologies, UK (ONT) for support with consumables and Vania Costa (ONT) for support with DNA extraction and library preparation for R10.4.1 samples. We also thank Megan Baxter for her input for future studies regarding *FN1* deletion.

This work was supported by the Edinburgh International Data Facility (EIDF) and the Data-Driven Innovation Programme at the University of Edinburgh.

This research was made possible through access to data in the National Genomic Research Library, which is managed by Genomics England Limited (a wholly owned company of the Department of Health and Social Care). The National Genomic Research Library holds data provided by patients and collected by the NHS as part of their care and data collected as part of their participation in research. The National Genomic Research Library is funded by the National Institute for Health Research and NHS England. The Wellcome Trust, Cancer Research UK and the Medical Research Council have also funded research infrastructure.

The views expressed are those of the authors and not necessarily those of the NHS, the NIHR or the Department of Health or Wellcome Trust.

## Supplementary Information

Additional File 1: Supplementary Tables 1 -7 and Supplementary Figures 1-7

Additional File 2: Supplementary Tables 8 and 9

