## Additional File 1 for "Detecting pathogenic structural variation in families with undiagnosed rare disease in a national genome project"

#### Supplementary Tables and Figures

This additional file contains supplementary tables 1-7 and supplementary figures 1-7.

### Supplementary Tables

#### Supplementary Table 1

| S.No. | Sample | Family Structure | Flowcells_used |
|---|---|---|---|
| 1 | Family_1_Father | Trio | R9+R10 |
| 2 | Family_1_Mother | Trio | R9+R10 |
| 3 | Family_1_Proband | Trio | R9+R10 |
| 4 | Family_2_Father | Trio | R9 |
| 5 | Family_2_Mother | Trio | R9 |
| 6 | Family_2_Proband | Trio | R9 |
| 7 | Family_3_Father | Trio | R9+R10 |
| 8 | Family_3_Mother | Trio | R9+R10 |
| 9 | Family_3_Proband | Trio | R9+R10 |
| 10 | Family_4_Father | Quad | R9+R10 |
| 11 | Family_4_Mother | Quad | R9+R10 |
| 12 | Family_4_Proband | Quad | R9+R10 |
| 13 | Family_4_Sibling | Quad | R10 |
| 14 | Family_5_Father | Trio | R9 |
| 15 | Family_5_Mother | Trio | R9 |
| 16 | Family_5_Proband | Trio | R9 |
| 17 | Family_6_Father | Trio | R9 |
| 18 | Family_6_Mother | Trio | R9 |
| 19 | Family_6_Proband | Trio | R9 |
| 20 | Family_7_Father | Trio | R9 |
| 21 | Family_7_Mother | Trio | R9 |
| 22 | Family_7_Proband | Trio | R9 |
| 23 | Family_8_Father | Quad | R9+R10 |
| 24 | Family_8_Mother | Quad | R9+R10 |
| 25 | Family_8_Proband | Quad | R9+R10 |
| 26 | Family_8_Sibling | Quad | R9+R10 |
| 27 | Family_9_Father | Trio | R9+R10 |
| 28 | Family_9_Mother | Trio | R9+R10 |
| 29 | Family_9_Proband | Trio | R9+R10 |
| 30 | Family_10_Father | Trio | R9+R10 |
| 31 | Family_10_Mother | Trio | R9+R10 |
| 32 | Family_10_Proband | Trio | R9+R10 |
| 33 | Family_11_Father | Trio | R9 |
| 34 | Family_11_Mother | Trio | R9 |
| 35 | Family_11_Proband | Trio | R9 |
| 36 | Family_12_Father | Trio | R9 |
| 37 | Family_12_Mother | Trio | R9 |
| 38 | Family_12_Proband | Trio | R9 |
| 39 | Family_13_Father | Trio | R9+R10 |
| 40 | Family_13_Mother | Trio | R9+R10 |
| 41 | Family_13_Proband | Trio | R9+R10 |
| 42 | Family_14_Father | Trio | R9 |
| 43 | Family_14_Mother | Trio | R9 |
| 44 | Family_14_Proband | Trio | R9 |
| 45 | Family_15_Father | Trio | R9+R10 |
| 46 | Family_15_Mother | Trio | R9+R10 |
| 47 | Family_15_Proband | Trio | R9+R10 |
| 48 | Family_16_Father | Trio | R9 |
| 49 | Family_16_Mother | Trio | R9 |
| 50 | Family_16_Proband | Trio | R9 |
| 51 | Family_17_Father | Trio | R9 |

| S.No. | Sample | Family Structure | Flowcells_used |
|---|---|---|---|
| 52 | Family_17_Mother | Trio | R9 |
| 53 | Family_17_Proband | Trio | R9 |
| 54 | Family_18_Father | Trio | R9+R10 |
| 55 | Family_18_Mother | Trio | R9+R10 |
| 56 | Family_18_Proband | Trio | R9+R10 |
| 57 | Family_19_Father | Trio | R9 |
| 58 | Family_19_Mother | Trio | R9 |
| 59 | Family_19_Proband | Trio | R9 |
| 60 | Family_20_Father | Trio | R9 |
| 61 | Family_20_Mother | Trio | R9 |
| 62 | Family_20_Proband | Trio | R9 |
| 63 | Family_21_Father | Trio | R9 |
| 64 | Family_21_Mother | Trio | R9 |
| 65 | Family_21_Proband | Trio | R9 |
| 66 | Family_22_Father | Trio | R9 |
| 67 | Family_22_Mother | Trio | R9 |
| 68 | Family_22_Proband | Trio | R9 |
| 69 | Family_23_Father | Trio | R9 |
| 70 | Family_23_Mother | Trio | R9 |
| 71 | Family_23_Proband | Trio | R9 |
| 72 | Family_24_Father | Trio | R9 |
| 73 | Family_24_Mother | Trio | R9 |
| 74 | Family_24_Proband | Trio | R9 |

**Supplementary Table 1: Information on the family structure and type of sequencing flowcell (R9 or R10 or R9+R10) used for n=74 samples. It should be noted that R9 is used as a short form for R9.4.1, and R10 is used as a short form for R10.4.1.**

#### Supplementary Table 2

| S.No. | Sample | Total bases_fastq | Number of reads_fastq | Median read quality_fastq | Median read length_fastq | Read length N50_fastq |
|---|---|---|---|---|---|---|
| 1 | Family_1_Father | 57824958580 | 3583850 | 14.1 | 16297 | 22597 |
| 2 | Family_1_Mother | 57271329301 | 3113415 | 14.2 | 18080 | 25401 |
| 3 | Family_1_Proband | 51355171007 | 2922528 | 14.1 | 16621 | 24765 |
| 4 | Family_2_Father | 46096538838 | 2379778 | 13.2 | 19787 | 27589 |
| 5 | Family_2_Mother | 49487226342 | 3852967 | 13.3 | 9783 | 22024 |
| 6 | Family_2_Proband | 55509431989 | 3320027 | 13.7 | 16338 | 22323 |
| 7 | Family_3_Father | 52306239408 | 3044049 | 14.4 | 16946 | 25008 |
| 8 | Family_3_Mother | 57495429746 | 3430311 | 13.9 | 15978 | 25032 |
| 9 | Family_3_Proband | 40343412192 | 2555879 | 13.1 | 16248 | 22222 |
| 10 | Family_4_Father | 54651526648 | 3415421 | 13.3 | 16648 | 22974 |
| 11 | Family_4_Mother | 35732033718 | 2313183 | 12.8 | 16246 | 22621 |
| 12 | Family_4_Proband | 46751150998 | 2900243 | 13.4 | 16382 | 23433 |
| 13 | Family_5_Father | 37191773204 | 3687972 | 13.6 | 6352 | 18639 |
| 14 | Family_5_Mother | 38832125403 | 3119048 | 12.9 | 9412 | 20476 |
| 15 | Family_5_Proband | 49614723469 | 3596343 | 14.4 | 14810 | 19992 |
| 16 | Family_6_Father | 61260899941 | 3841540 | 14.4 | 16455 | 20999 |
| 17 | Family_6_Mother | 66201070662 | 3454849 | 14.8 | 18800 | 24958 |
| 18 | Family_6_Proband | 57442646360 | 3461929 | 14 | 16663 | 21618 |
| 19 | Family_7_Father | 44008423302 | 3767207 | 14.3 | 9414 | 18387 |
| 20 | Family_7_Mother | 66361603960 | 5594691 | 14.6 | 10874 | 17493 |
| 21 | Family_7_Proband | 64194031918 | 4112975 | 14.5 | 15975 | 21276 |
| 22 | Family_8_Father | 49436127391 | 2538851 | 13.1 | 20397 | 26377 |
| 23 | Family_8_Mother | 52054260828 | 2511657 | 12.7 | 21208 | 27811 |
| 24 | Family_8_Proband | 45272212284 | 2907896 | 13.1 | 13542 | 25094 |
| 25 | Family_8_Sibling | 37835615628 | 1973096 | 12.8 | 19562 | 26275 |
| 26 | Family_9_Father | 56502627816 | 3895955 | 13.5 | 12510 | 22915 |
| 27 | Family_9_Mother | 39876539241 | 2381218 | 13.6 | 16384 | 25368 |
| 28 | Family_9_Proband | 35355007544 | 1975369 | 13.5 | 17882 | 24623 |
| 29 | Family_10_Father | 75287504146 | 4420209 | 15 | 17358 | 23781 |
| 30 | Family_10_Mother | 42464987854 | 2685381 | 14.2 | 15268 | 23475 |
| 31 | Family_10_Proband | 64150375839 | 5456240 | 14.4 | 9643 | 18003 |
| 32 | Family_11_Father | 50560975645 | 2630563 | 12.9 | 19214 | 26008 |
| 33 | Family_11_Mother | 46395572179 | 2522915 | 13.5 | 18167 | 25873 |
| 34 | Family_11_Proband | 99434800280 | 5455164 | 14.4 | 19498 | 24383 |
| 35 | Family_12_Father | 60563989980 | 5400441 | 13.3 | 9324 | 17292 |
| 36 | Family_12_Mother | 46110898995 | 3217641 | 13.9 | 12329 | 22068 |
| 37 | Family_12_Proband | 43942774271 | 3354679 | 12.9 | 10761 | 20675 |
| 38 | Family_13_Father | 39450608571 | 2569297 | 12.9 | 16690 | 23201 |
| 39 | Family_13_Mother | 59855710767 | 3358261 | 13 | 18201 | 25570 |
| 40 | Family_13_Proband | 56321986536 | 3781359 | 14.4 | 15380 | 21286 |
| 41 | Family_14_Father | 64165629861 | 3676689 | 13.9 | 17479 | 24074 |
| 42 | Family_14_Mother | 62109434023 | 3562701 | 13.7 | 17142 | 24529 |
| 43 | Family_14_Proband | 62882866442 | 3675816 | 14.2 | 13003 | 30421 |
| 44 | Family_15_Father | 49115705659 | 3574146 | 13.5 | 11505 | 21647 |
| 45 | Family_15_Mother | 47183016415 | 2929562 | 14.1 | 14713 | 24169 |
| 46 | Family_15_Proband | 56846993178 | 3417612 | 14.2 | 15909 | 23900 |
| 47 | Family_16_Father | 48851317869 | 3022076 | 14.5 | 15652 | 23552 |
| 48 | Family_16_Mother | 41368469685 | 2904171 | 14.6 | 12005 | 21963 |
| 49 | Family_16_Proband | 39299754367 | 3325982 | 14.3 | 9302 | 19687 |
| 50 | Family_17_Father | 57824611229 | 3169820 | 13.5 | 18241 | 24718 |
| 51 | Family_17_Mother | 68015190327 | 3855169 | 14.3 | 18494 | 23886 |
| 52 | Family_17_Proband | 63006122934 | 3200453 | 14.3 | 20189 | 26909 |
| 53 | Family_18_Father | 53672645242 | 2678845 | 13.2 | 20755 | 24831 |

| S.No. | Sample | Total bases_fastq | Number of reads_fastq | Median read quality_fastq | Median read length_fastq | Read length N50_fastq |
|---|---|---|---|---|---|---|
| 54 | Family_18_Mother | 39863232261 | 2134263 | 12.6 | 19623 | 24954 |
| 55 | Family_18_Proband | 55070814543 | 2784123 | 12.5 | 20811 | 25625 |
| 56 | Family_19_Father | 70997662658 | 4197967 | 14.1 | 16373 | 23981 |
| 57 | Family_19_Mother | 55935351112 | 2934575 | 14.2 | 19008 | 25864 |
| 58 | Family_19_Proband | 56241787237 | 3604131 | 13.7 | 15747 | 22113 |
| 59 | Family_20_Father | 39663270377 | 2302819 | 13 | 17190 | 24917 |
| 60 | Family_20_Mother | 53486426275 | 3486750 | 13 | 13733 | 24024 |
| 61 | Family_20_Proband | 55115177963 | 2654939 | 12.7 | 21621 | 27227 |
| 62 | Family_21_Father | 32548002238 | 2104943 | 13.6 | 14276 | 22885 |
| 63 | Family_21_Mother | 31978382392 | 2301489 | 13.7 | 11620 | 21486 |
| 64 | Family_21_Proband | 42490345930 | 3074226 | 14.2 | 11854 | 21529 |
| 65 | Family_22_Father | 32038063331 | 2489070 | 13.1 | 11223 | 19234 |
| 66 | Family_22_Mother | 36986979119 | 2571231 | 14 | 12526 | 21812 |
| 67 | Family_22_Proband | 48867187400 | 4371072 | 13.7 | 9591 | 16961 |
| 68 | Family_23_Father | 43849477723 | 3148263 | 14.2 | 12289 | 21008 |
| 69 | Family_23_Mother | 41922733398 | 3043747 | 14.6 | 12283 | 20224 |
| 70 | Family_23_Proband | 31726184177 | 2744711 | 14 | 8626 | 19009 |
| 71 | Family_24_Father | 50474719033 | 2829944 | 14.1 | 17779 | 24404 |
| 72 | Family_24_Mother | 51677739739 | 3090901 | 14.7 | 16231 | 24069 |
| 73 | Family_24_Proband | 61121920468 | 3344186 | 12.8 | 18651 | 23588 |

**Supplementary Table 2: Raw FASTQ QC metrics for n=73 samples sequenced using R9.4.1 flowcell**

#### Supplementary Table 3

| S.No. | Sample | Total bases_fastq | Number of reads_fastq | Median read quality_fastq | Median read length_fastq | Read length N50_fastq |
|---|---|---|---|---|---|---|
| 1 | Family_1_Father_trimmed_filtered | 56854148578 | 3250029 | 14.2 | 17458 | 22663 |
| 2 | Family_1_Mother_trimmed_filtered | 56424626174 | 2854592 | 14.3 | 19248 | 25487 |
| 3 | Family_1_Proband_trimmed_filtered | 50478765952 | 2663746 | 14.2 | 17859 | 24883 |
| 4 | Family_2_Father_trimmed_filtered | 41658436577 | 1878850 | 13.5 | 22225 | 27783 |
| 5 | Family_2_Mother_trimmed_filtered | 44539940744 | 2889624 | 13.7 | 14098 | 22367 |
| 6 | Family_2_Proband_trimmed_filtered | 50703698996 | 2679353 | 14 | 17983 | 22489 |
| 7 | Family_3_Father_trimmed_filtered | 50898014816 | 2575892 | 14.5 | 19135 | 25246 |
| 8 | Family_3_Mother_trimmed_filtered | 55955974371 | 2913490 | 14 | 18168 | 25309 |
| 9 | Family_3_Proband_trimmed_filtered | 39356680425 | 2211939 | 13.2 | 17691 | 22385 |
| 10 | Family_4_Father_trimmed_filtered | 50020493482 | 2807512 | 13.6 | 18126 | 23045 |
| 11 | Family_4_Mother_trimmed_filtered | 31385372686 | 1767597 | 13.1 | 18129 | 22714 |
| 12 | Family_4_Proband_trimmed_filtered | 43347874751 | 2399068 | 13.7 | 18064 | 23523 |
| 13 | Family_5_Father_trimmed_filtered | 36124220182 | 3150826 | 13.6 | 8139 | 18929 |
| 14 | Family_5_Mother_trimmed_filtered | 37787149948 | 2741179 | 13 | 11281 | 20731 |
| 15 | Family_5_Proband_trimmed_filtered | 47995574602 | 3056558 | 14.5 | 17050 | 20135 |
| 16 | Family_6_Father_trimmed_filtered | 59997849342 | 3442912 | 14.5 | 17406 | 21082 |
| 17 | Family_6_Mother_trimmed_filtered | 65062249925 | 3136132 | 14.8 | 19886 | 25071 |
| 18 | Family_6_Proband_trimmed_filtered | 56439712574 | 3158925 | 14.1 | 17500 | 21701 |
| 19 | Family_7_Father_trimmed_filtered | 42984937125 | 3354236 | 14.4 | 10971 | 18546 |
| 20 | Family_7_Mother_trimmed_filtered | 64865978960 | 5029721 | 14.7 | 12593 | 17570 |
| 21 | Family_7_Proband_trimmed_filtered | 63195023703 | 3790226 | 14.6 | 16693 | 21335 |
| 22 | Family_8_Father_trimmed_filtered | 44055668791 | 1997311 | 13.4 | 22163 | 26505 |
| 23 | Family_8_Mother_trimmed_filtered | 46149029702 | 1996607 | 13.2 | 22931 | 27959 |
| 24 | Family_8_Proband_trimmed_filtered | 40129272945 | 2247184 | 13.5 | 17278.5 | 25154 |
| 25 | Family_8_Sibling_trimmed_filtered | 33117992039 | 1547570 | 13.3 | 21207 | 26393 |
| 26 | Family_9_Father_trimmed_filtered | 54782392857 | 3272231 | 13.6 | 15754 | 23231 |
| 27 | Family_9_Mother_trimmed_filtered | 38854489665 | 2024454 | 13.7 | 19126 | 25576 |
| 28 | Family_9_Proband_trimmed_filtered | 34670023142 | 1757808 | 13.6 | 19307 | 24751 |
| 29 | Family_10_Father_trimmed_filtered | 73547713109 | 3880045 | 15.1 | 18934 | 23930 |
| 30 | Family_10_Mother_trimmed_filtered | 41500497300 | 2356029 | 14.3 | 17072 | 23664 |
| 31 | Family_10_Proband_trimmed_filtered | 63069725484 | 4993565 | 14.5 | 10792 | 18092 |
| 32 | Family_11_Father_trimmed_filtered | 49614620612 | 2373918 | 13 | 20499 | 26106 |
| 33 | Family_11_Mother_trimmed_filtered | 45343395245 | 2233995 | 13.5 | 19846 | 26016 |
| 34 | Family_11_Proband_trimmed_filtered | 96876392828 | 4656766 | 14.5 | 21056 | 24501 |
| 35 | Family_12_Father_trimmed_filtered | 59520305055 | 4915257 | 13.4 | 10581 | 17321 |
| 36 | Family_12_Mother_trimmed_filtered | 45183065545 | 2898720 | 14 | 14236 | 22200 |
| 37 | Family_12_Proband_trimmed_filtered | 43196922066 | 3056230 | 13 | 12193 | 20791 |
| 38 | Family_13_Father_trimmed_filtered | 34774533702 | 1974511 | 13.4 | 18534 | 23264 |
| 39 | Family_13_Mother_trimmed_filtered | 53148368297 | 2613218 | 13.4 | 20301 | 25750 |
| 40 | Family_13_Proband_trimmed_filtered | 54975884430 | 3352932 | 14.5 | 16849 | 21386 |
| 41 | Family_14_Father_trimmed_filtered | 62861702077 | 3272955 | 14 | 18845 | 24213 |
| 42 | Family_14_Mother_trimmed_filtered | 60780118521 | 3138987 | 13.8 | 18801 | 24687 |
| 43 | Family_14_Proband_trimmed_filtered | 60732890101 | 2971490 | 14.3 | 18268 | 30948 |
| 44 | Family_15_Father_trimmed_filtered | 48249702659 | 3206200 | 13.6 | 13486 | 21775 |
| 45 | Family_15_Mother_trimmed_filtered | 46148037990 | 2608143 | 14.2 | 16838 | 24345 |
| 46 | Family_15_Proband_trimmed_filtered | 56028744190 | 3126001 | 14.2 | 17098 | 23999 |
| 47 | Family_16_Father_trimmed_filtered | 47900769629 | 2733948 | 14.6 | 17227 | 23667 |
| 48 | Family_16_Mother_trimmed_filtered | 40344171230 | 2572853 | 14.7 | 14189 | 22183 |
| 49 | Family_16_Proband_trimmed_filtered | 37897007217 | 2761842 | 14.5 | 12577 | 19903 |
| 50 | Family_17_Father_trimmed_filtered | 56790709827 | 2869382 | 13.6 | 19451 | 24803 |
| 51 | Family_17_Mother_trimmed_filtered | 66698773396 | 3462279 | 14.4 | 19621 | 23956 |
| 52 | Family_17_Proband_trimmed_filtered | 61831296145 | 2848265 | 14.4 | 21733 | 27024 |
| 53 | Family_18_Father_trimmed_filtered | 48249310158 | 2205424 | 13.6 | 21675 | 24871 |

| S.No. | Sample | Total bases_fastq | Number of reads_fastq | Median read quality_fastq | Median read length_fastq | Read length N50_fastq |
|---|---|---|---|---|---|---|
| 54 | Family_18_Mother_trimmed_filtered | 34620173724 | 1670708 | 13.1 | 20929 | 25013 |
| 55 | Family_18_Proband_trimmed_filtered | 48399950584 | 2163324 | 12.9 | 22284 | 25721 |
| 56 | Family_19_Father_trimmed_filtered | 69621601251 | 3778204 | 14.1 | 17701 | 24115 |
| 57 | Family_19_Mother_trimmed_filtered | 54165290239 | 2460590 | 14.2 | 21156 | 26130 |
| 58 | Family_19_Proband_trimmed_filtered | 54799450653 | 3129089 | 13.8 | 17269 | 22289 |
| 59 | Family_20_Father_trimmed_filtered | 34973811584 | 1813278 | 13.5 | 18957 | 25012 |
| 60 | Family_20_Mother_trimmed_filtered | 47945259094 | 2820582 | 13.3 | 16390 | 24004 |
| 61 | Family_20_Proband_trimmed_filtered | 48265646136 | 2063144 | 13.2 | 23277 | 27259 |
| 62 | Family_21_Father_trimmed_filtered | 32045222176 | 1920651 | 13.7 | 15849 | 22994 |
| 63 | Family_21_Mother_trimmed_filtered | 31466030348 | 2091790 | 13.7 | 13262 | 21616 |
| 64 | Family_21_Proband_trimmed_filtered | 41409731402 | 2707614 | 14.3 | 14337 | 21702 |
| 65 | Family_22_Father_trimmed_filtered | 31294606788 | 2232942 | 13.2 | 12958 | 19389 |
| 66 | Family_22_Mother_trimmed_filtered | 36181971400 | 2307130 | 14.1 | 14432 | 21977 |
| 67 | Family_22_Proband_trimmed_filtered | 47798453916 | 3939154 | 13.8 | 11023 | 17051 |
| 68 | Family_23_Father_trimmed_filtered | 42782776036 | 2784641 | 14.3 | 14395 | 21201 |
| 69 | Family_23_Mother_trimmed_filtered | 41091016347 | 2765773 | 14.7 | 13813 | 20354 |
| 70 | Family_23_Proband_trimmed_filtered | 30990172494 | 2435602 | 14.1 | 10159 | 19195 |
| 71 | Family_24_Father_trimmed_filtered | 49545876239 | 2567664 | 14.2 | 18958 | 24491 |
| 72 | Family_24_Mother_trimmed_filtered | 50567515368 | 2736597 | 14.8 | 17741 | 24230 |
| 73 | Family_24_Proband_trimmed_filtered | 52497674732 | 2556057 | 13.2 | 20031 | 23523 |

**Supplementary Table 3: Trimmed and filtered FASTQ QC metrics for n=73 samples sequenced using R9.4.1 flowcell**

#### Supplementary Table 4

| S.No. | Sample | Total bases_fastq | Number of reads_fastq | Median read quality_fastq | Median read length_fastq | Read length N50_fastq |
|---|---|---|---|---|---|---|
| 1 | Family_1_Father | 103239559381 | 19072681 | 18.3 | 2416 | 12660 |
| 2 | Family_1_Mother | 94971191088 | 15192865 | 18.6 | 2940 | 13934 |
| 3 | Family_1_Proband | 92589215167 | 12085760 | 18.9 | 3931 | 15948 |
| 4 | Family_3_Father | 113123998342 | 21676581 | 18.4 | 3161 | 8918 |
| 5 | Family_3_Mother | 116301918708 | 18701934 | 18.7 | 3545 | 11289 |
| 6 | Family_3_Proband | 87072186608 | 15485693 | 18.6 | 3212 | 10229 |
| 7 | Family_4_Sibling | 109965766880 | 23422189 | 18.4 | 2573 | 8633 |
| 8 | Family_4_Father | 113228877921 | 36789519 | 17.4 | 1588 | 5733 |
| 9 | Family_4_Mother | 141506552578 | 43825712 | 18.2 | 2201 | 4766 |
| 10 | Family_4_Proband | 132832199693 | 38925450 | 17.7 | 1767 | 6444 |
| 11 | Family_8_Father | 118825762491 | 19717271 | 18.5 | 2937 | 13068 |
| 12 | Family_8_Mother | 85999081935 | 17628250 | 18.1 | 2169 | 11659 |
| 13 | Family_8_Proband | 118997710924 | 39550354 | 17.5 | 1291 | 6936 |
| 14 | Family_8_Sibling | 108598671107 | 38548894 | 17.5 | 1246 | 6342 |
| 15 | Family_9_Father | 85619101476 | 19266224 | 18 | 2115 | 9320 |
| 16 | Family_9_Mother | 94368142329 | 18562800 | 18.3 | 2438 | 10744 |
| 17 | Family_9_Proband | 97864666827 | 18342286 | 18.5 | 2654 | 10793 |
| 18 | Family_10_Father | 70549900680 | 18185887 | 17.4 | 1725 | 9269 |
| 19 | Family_10_Mother | 82888052802 | 18577981 | 17.7 | 1829 | 11691 |
| 20 | Family_10_Proband | 63104207596 | 17216211 | 17.3 | 1419 | 10367 |
| 21 | Family_13_Father | 114848402635 | 45816218 | 17.5 | 1402 | 4223 |
| 22 | Family_13_Mother | 114075012024 | 30447203 | 18.2 | 2440 | 5827 |
| 23 | Family_13_Proband | 88848902151 | 14363249 | 18.6 | 3485 | 11128 |
| 24 | Family_15_Father | 140253530958 | 53919664 | 18 | 1839 | 3738 |
| 25 | Family_15_Mother | 97244846577 | 13683618 | 18.8 | 4258 | 12029 |
| 26 | Family_15_Proband | 97994614810 | 13926547 | 18.9 | 4267 | 11665 |
| 27 | Family_18_Father | 80516750327 | 11369179 | 18.9 | 4031 | 13203 |
| 28 | Family_18_Mother | 109695720308 | 15472894 | 18.8 | 4070 | 13083 |
| 29 | Family_18_Proband | 98692514416 | 17119029 | 18.3 | 2972 | 11625 |

**Supplementary Table 4: Raw FASTQ QC metrics for n=29 samples sequenced using R10.4.1 flowcell**

#### Supplementary Table 5

| S.No. | Sample | Total bases_fastq | Number of reads_fastq | Median read quality_fastq | Median read length_fastq | Read length N50_fastq |
|---|---|---|---|---|---|---|
| 1 | Family_1_Father_trimmed_filtered | 99244277297 | 13811178 | 20 | 3713 | 13467 |
| 2 | Family_1_Mother_trimmed_filtered | 92199304222 | 11837330 | 20 | 4057 | 14553 |
| 3 | Family_1_Proband_trimmed_filtered | 90951538826 | 10353085 | 20 | 4655 | 16362 |
| 4 | Family_3_Father_trimmed_filtered | 109416195762 | 17629893 | 19.8 | 4009 | 9186 |
| 5 | Family_3_Mother_trimmed_filtered | 113396319373 | 15601139 | 19.9 | 4400 | 11575 |
| 6 | Family_3_Proband_trimmed_filtered | 84417228112 | 12475551 | 19.9 | 4225 | 10522 |
| 7 | Family_4_Sibling_trimmed_filtered | 105342025398 | 18091953 | 20 | 3500 | 9062 |
| 8 | Family_4_Father_trimmed_filtered | 103577498415 | 23252450 | 19.6 | 2660 | 6416 |
| 9 | Family_4_Mother_trimmed_filtered | 132776903281 | 33955879 | 19.9 | 2774 | 4999 |
| 10 | Family_4_Proband_trimmed_filtered | 123415607156 | 26496179 | 19.7 | 2720 | 7075 |
| 11 | Family_8_Father_trimmed_filtered | 115014710388 | 14882185 | 20 | 4292 | 13625 |
| 12 | Family_8_Mother_trimmed_filtered | 82178628692 | 12482344 | 19.9 | 3305 | 12570 |
| 13 | Family_8_Proband_trimmed_filtered | 106941920650 | 22673733 | 19.9 | 2334 | 8367 |
| 14 | Family_8_Sibling_trimmed_filtered | 96594896072 | 21844199 | 19.9 | 2230 | 7795 |
| 15 | Family_9_Father_trimmed_filtered | 81317020310 | 13890816 | 19.8 | 3131 | 10080 |
| 16 | Family_9_Mother_trimmed_filtered | 90542553942 | 13817885 | 19.9 | 3506 | 11434 |
| 17 | Family_9_Proband_trimmed_filtered | 94327855305 | 14229215 | 20 | 3601 | 11374 |
| 18 | Family_10_Father_trimmed_filtered | 66223161392 | 12180375 | 19.4 | 2664 | 10457 |
| 19 | Family_10_Mother_trimmed_filtered | 78603953368 | 12672873 | 19.6 | 2909 | 12748 |
| 20 | Family_10_Proband_trimmed_filtered | 58518622374 | 10493016 | 19.5 | 2515 | 11748 |
| 21 | Family_13_Father_trimmed_filtered | 101630406215 | 27691586 | 19.8 | 2281 | 4903 |
| 22 | Family_13_Mother_trimmed_filtered | 108131854219 | 23644553 | 19.8 | 3148 | 6075 |
| 23 | Family_13_Proband_trimmed_filtered | 86751020653 | 12365847 | 19.7 | 4116 | 11426 |
| 24 | Family_15_Father_trimmed_filtered | 127815639533 | 38703513 | 19.9 | 2456 | 3992 |
| 25 | Family_15_Mother_trimmed_filtered | 95632859475 | 12303919 | 19.8 | 4730 | 12221 |
| 26 | Family_15_Proband_trimmed_filtered | 96397486964 | 12528494 | 19.8 | 4722 | 11843 |
| 27 | Family_18_Father_trimmed_filtered | 78969944307 | 9847631 | 20 | 4745 | 13470 |
| 28 | Family_18_Mother_trimmed_filtered | 107595012823 | 13547749 | 19.9 | 4783 | 13314 |
| 29 | Family_18_Proband_trimmed_filtered | 95595580967 | 13557441 | 19.7 | 3991 | 12103 |

**Supplementary Table 5: Trimmed and filtered FASTQ QC metrics for n=29 samples sequenced using R10.4.1 flowcell**

#### Supplementary Table 6

| S.No. | Sample | Mean identity % (NanoComp) | Median identity % (NanoComp) | Total reads (Samtools) | Mapped reads (Samtools) | Mapped reads % (Samtools) | Median Coverage (Qualimap) | Mean Coverage (Qualimap) | Flowcells_used |
|---|---|---|---|---|---|---|---|---|---|
| 1 | Family_1_Father | 96.2 | 98.3 | 18314879 | 18308052 | 99.96 | 50X | 50.8X | R9+R10 |
| 2 | Family_1_Mother | 96.2 | 98.3 | 15719038 | 15713009 | 99.96 | 47X | 48.3X | R9+R10 |
| 3 | Family_1_Proband | 96 | 98.2 | 14035784 | 14031608 | 99.97 | 45X | 46.0X | R9+R10 |
| 4 | Family_2_Father | 91.7 | 94 | 2064495 | 2062933 | 99.92 | 13X | 13.7X | R9 |
| 5 | Family_2_Mother | 92.6 | 94.4 | 3031216 | 3028888 | 99.92 | 14X | 14.6X | R9 |
| 6 | Family_2_Proband | 92.5 | 94.7 | 2860574 | 2855119 | 99.81 | 16X | 16.6X | R9 |
| 7 | Family_3_Father | 96.2 | 98.4 | 21464390 | 21455717 | 99.96 | 51X | 52.2X | R9+R10 |
| 8 | Family_3_Mother | 96.2 | 98.4 | 19835837 | 19827620 | 99.96 | 54X | 55.0X | R9+R10 |
| 9 | Family_3_Proband | 96 | 98.4 | 15893705 | 15890038 | 99.98 | 40X | 40.3X | R9+R10 |
| 10 | Family_4_Father | 96.6 | 98.4 | 27527865 | 27512633 | 99.94 | 49X | 50.0X | R9+R10 |
| 11 | Family_4_Mother | 97.1 | 98.6 | 37529232 | 37512486 | 99.96 | 53X | 53.3X | R9+R10 |
| 12 | Family_4_Proband | 96.9 | 98.5 | 30304566 | 30289547 | 99.95 | 53X | 54.2X | R9+R10 |
| 13 | Family_4_Sibling | 97.2 | 98.7 | 19136961 | 19133588 | 99.98 | 34X | 34.2X | R10 |
| 14 | Family_5_Father | 92.8 | 94.7 | 3422263 | 3421612 | 99.98 | 11X | 11.8X | R9 |
| 15 | Family_5_Mother | 92 | 93.8 | 3030898 | 3030331 | 99.98 | 12X | 12.4X | R9 |
| 16 | Family_5_Proband | 93.3 | 95.5 | 3365261 | 3364313 | 99.97 | 15X | 15.6X | R9 |
| 17 | Family_6_Father | 93.3 | 95.5 | 3797483 | 3797012 | 99.99 | 19X | 19.5X | R9 |
| 18 | Family_6_Mother | 93.5 | 95.8 | 3476819 | 3476429 | 99.99 | 21X | 21.2X | R9 |
| 19 | Family_6_Proband | 92.6 | 95.1 | 3656595 | 3656184 | 99.99 | 18X | 18.4X | R9 |
| 20 | Family_7_Father | 93.4 | 95.5 | 3648401 | 3648041 | 99.99 | 13X | 14.0X | R9 |
| 21 | Family_7_Mother | 93.7 | 95.8 | 5452299 | 5451883 | 99.99 | 20X | 21.1X | R9 |
| 22 | Family_7_Proband | 93.4 | 95.6 | 4120097 | 4118623 | 99.96 | 20X | 20.6X | R9 |
| 23 | Family_8_Father | 96.1 | 98.4 | 18048236 | 18043580 | 99.97 | 51X | 51.9X | R9+R10 |
| 24 | Family_8_Mother | 96.2 | 98.4 | 15418752 | 15412982 | 99.96 | 41X | 41.7X | R9+R10 |
| 25 | Family_8_Proband | 96.7 | 98.5 | 27234290 | 27216909 | 99.94 | 47X | 47.8X | R9+R10 |
| 26 | Family_8_Sibling | 97 | 98.6 | 25648512 | 25639937 | 99.97 | 42X | 42.1X | R9+R10 |
| 27 | Family_9_Father | 96.1 | 98.3 | 18617794 | 18615357 | 99.99 | 44X | 44.3X | R9+R10 |
| 28 | Family_9_Mother | 96.4 | 98.4 | 16937467 | 16934906 | 99.98 | 41X | 42.1X | R9+R10 |
| 29 | Family_9_Proband | 96.5 | 98.5 | 17147896 | 17145822 | 99.99 | 41X | 42.0X | R9+R10 |
| 30 | Family_10_Father | 96.2 | 98 | 17526801 | 17523712 | 99.98 | 45X | 45.4X | R9+R10 |
| 31 | Family_10_Mother | 96.5 | 98.3 | 16300312 | 16294585 | 99.96 | 39X | 39.0X | R9+R10 |
| 32 | Family_10_Proband | 96.1 | 97.9 | 16811956 | 16809155 | 99.98 | 39X | 39.4X | R9+R10 |
| 33 | Family_11_Father | 92 | 93.8 | 2617201 | 2616076 | 99.96 | 16X | 16.2X | R9 |
| 34 | Family_11_Mother | 92.5 | 94.5 | 2464846 | 2464453 | 99.98 | 14X | 14.8X | R9 |
| 35 | Family_11_Proband | 93.3 | 95.6 | 5150019 | 5146615 | 99.93 | 31X | 31.6X | R9 |
| 36 | Family_12_Father | 92.3 | 94.3 | 5512826 | 5498734 | 99.74 | 19X | 19.4X | R9 |
| 37 | Family_12_Mother | 92.9 | 95 | 3217694 | 3216053 | 99.95 | 14X | 14.7X | R9 |
| 38 | Family_12_Proband | 92 | 93.8 | 3396476 | 3394809 | 99.95 | 13X | 14.1X | R9 |
| 39 | Family_13_Father | 97.1 | 98.5 | 32782911 | 32775745 | 99.98 | 44X | 44.3X | R9+R10 |
| 40 | Family_13_Mother | 96.6 | 98.5 | 27417495 | 27412817 | 99.98 | 51X | 52.6X | R9+R10 |
| 41 | Family_13_Proband | 96.2 | 98.2 | 16730367 | 16714821 | 99.91 | 45X | 46.1X | R9+R10 |
| 42 | Family_14_Father | 92.9 | 95 | 3606948 | 3606643 | 99.99 | 20X | 20.5X | R9 |
| 43 | Family_14_Mother | 92.8 | 94.8 | 3422220 | 3421315 | 99.97 | 19X | 19.9X | R9 |
| 44 | Family_14_Proband | 93 | 95.3 | 3381700 | 3380936 | 99.98 | 19X | 19.8X | R9 |
| 45 | Family_15_Father | 96.9 | 98.5 | 45327014 | 45310727 | 99.96 | 57X | 57.2X | R9+R10 |
| 46 | Family_15_Mother | 96.2 | 98.3 | 15857330 | 15855579 | 99.99 | 45X | 46.1X | R9+R10 |
| 47 | Family_15_Proband | 96.2 | 98.3 | 16574436 | 16568480 | 99.96 | 49X | 49.6X | R9+R10 |
| 48 | Family_16_Father | 93.2 | 95.5 | 3077239 | 3076298 | 99.97 | 15X | 15.6X | R9 |
| 49 | Family_16_Mother | 93.2 | 95.6 | 2911772 | 2911329 | 99.98 | 12X | 13.2X | R9 |
| 50 | Family_16_Proband | 93.3 | 95.6 | 3049156 | 3000506 | 98.4 | 12X | 12.3X | R9 |
| 51 | Family_17_Father | 92.5 | 94.5 | 3176694 | 3175855 | 99.97 | 18X | 18.6X | R9 |
| 52 | Family_17_Mother | 93.4 | 95.4 | 3806771 | 3806303 | 99.99 | 21X | 21.7X | R9 |
| 53 | Family_17_Proband | 93.2 | 95.4 | 3124850 | 3124457 | 99.99 | 20X | 20.2X | R9 |
| 54 | Family_18_Father | 96 | 98.3 | 12791216 | 12790141 | 99.99 | 41X | 41.4X | R9+R10 |
| 55 | Family_18_Mother | 96.3 | 98.5 | 16018349 | 16016196 | 99.99 | 45X | 46.4X | R9+R10 |
| 56 | Family_18_Proband | 96.2 | 98.4 | 16725742 | 16722938 | 99.98 | 46X | 46.9X | R9+R10 |
| 57 | Family_19_Father | 93 | 95.2 | 4207278 | 4206646 | 99.98 | 22X | 22.7X | R9 |
| 58 | Family_19_Mother | 93.1 | 95.4 | 2773513 | 2773021 | 99.98 | 17X | 17.7X | R9 |

| S.No. | Sample | Mean identity % (NanoComp) | Median identity % (NanoComp) | Total reads (Samtools) | Mapped reads (Samtools) | Mapped reads % (Samtools) | Median Coverage (Qualimap) | Mean Coverage (Qualimap) | Flowcells_used |
|---|---|---|---|---|---|---|---|---|---|
| 59 | Family_19_Proband | 92.8 | 94.8 | 3415555 | 3415226 | 99.99 | 17X | 17.8X | R9 |
| 60 | Family_20_Father | 92.2 | 94.1 | 1947988 | 1947750 | 99.99 | 11X | 11.5X | R9 |
| 61 | Family_20_Mother | 92.3 | 94.1 | 3042188 | 3030918 | 99.63 | 15X | 15.7X | R9 |
| 62 | Family_20_Proband | 91.9 | 93.7 | 2216423 | 2215623 | 99.96 | 15X | 15.8X | R9 |
| 63 | Family_21_Father | 92.5 | 94.7 | 2107022 | 2103874 | 99.85 | 10X | 10.5X | R9 |
| 64 | Family_21_Mother | 92.8 | 94.8 | 2261860 | 2258550 | 99.85 | 10X | 10.3X | R9 |
| 65 | Family_21_Proband | 93.1 | 95.4 | 3008432 | 3007626 | 99.97 | 13X | 13.5X | R9 |
| 66 | Family_22_Father | 92.2 | 94.1 | 2444692 | 2444337 | 99.99 | 10X | 10.2X | R9 |
| 67 | Family_22_Mother | 93.2 | 95.2 | 2546337 | 2546061 | 99.99 | 11X | 11.8X | R9 |
| 68 | Family_22_Proband | 92.9 | 94.9 | 4342157 | 4285057 | 98.68 | 15X | 15.4X | R9 |
| 69 | Family_23_Father | 93.2 | 95.3 | 3069016 | 3068313 | 99.98 | 13X | 13.9X | R9 |
| 70 | Family_23_Mother | 93.6 | 95.8 | 3039069 | 3038654 | 99.99 | 13X | 13.4X | R9 |
| 71 | Family_23_Proband | 93.4 | 95.3 | 2627117 | 2621316 | 99.78 | 10X | 10.1X | R9 |
| 72 | Family_24_Father | 93 | 95.2 | 2847721 | 2847073 | 99.98 | 16X | 16.2X | R9 |
| 73 | Family_24_Mother | 93.8 | 95.9 | 2989360 | 2988830 | 99.98 | 16X | 16.5X | R9 |
| 74 | Family_24_Proband | 91.5 | 93.6 | 2820434 | 2819417 | 99.96 | 16X | 17.2X | R9 |

**Supplementary Table 6: BAM file statistics for n=74 samples. Statistics calculated by three tools: NanoComp, samtools flagstat and Qualimap bamqc. The last columns tells if the sample was sequenced using only R9.4.1 flowcell (n=45) or using only R10.4.1 flowcell (n=1) or using both R9.4.1 and R10.4.1 flowcells (n=28).**

#### Supplementary Table 7

| S.No. | Family_ID | Total_SVs | INS | DEL | DUP | INV | TRA |
|---|---|---|---|---|---|---|---|
| 1 | Family_1 | 23024 | 12418 | 10087 | 17 | 37 | 465 |
| 2 | Family_2 | 23527 | 12709 | 10317 | 18 | 39 | 444 |
| 3 | Family_3 | 23403 | 12550 | 10265 | 24 | 49 | 515 |
| 4 | Family_4 | 24276 | 12973 | 10542 | 37 | 45 | 679 |
| 5 | Family_5 | 23869 | 12790 | 10425 | 23 | 52 | 579 |
| 6 | Family_6 | 23809 | 12898 | 10378 | 16 | 56 | 461 |
| 7 | Family_7 | 23930 | 12780 | 10549 | 21 | 46 | 534 |
| 8 | Family_8 | 24001 | 12853 | 10471 | 29 | 39 | 609 |
| 9 | Family_9 | 23655 | 12717 | 10369 | 27 | 35 | 507 |
| 10 | Family_10 | 23633 | 12812 | 10243 | 33 | 42 | 503 |
| 11 | Family_11 | 23374 | 12577 | 10370 | 21 | 45 | 361 |
| 12 | Family_12 | 24656 | 13248 | 10801 | 21 | 44 | 542 |
| 13 | Family_13 | 23499 | 12610 | 10247 | 31 | 44 | 567 |
| 14 | Family_14 | 25009 | 13489 | 11080 | 17 | 44 | 379 |
| 15 | Family_15 | 23650 | 12663 | 10280 | 29 | 44 | 634 |
| 16 | Family_16 | 24012 | 12933 | 10467 | 26 | 57 | 529 |
| 17 | Family_17 | 23797 | 12834 | 10535 | 13 | 38 | 377 |
| 18 | Family_18 | 23273 | 12577 | 10138 | 23 | 38 | 497 |
| 19 | Family_19 | 23863 | 12935 | 10469 | 21 | 44 | 394 |
| 20 | Family_20 | 23819 | 12724 | 10608 | 23 | 45 | 419 |
| 21 | Family_21 | 23917 | 12811 | 10452 | 33 | 50 | 571 |
| 22 | Family_22 | 24001 | 12859 | 10453 | 28 | 52 | 609 |
| 23 | Family_23 | 24041 | 12877 | 10527 | 23 | 47 | 567 |
| 24 | Family_24 | 23609 | 12709 | 10424 | 25 | 33 | 418 |

**Supplementary Table 7: SV statistics of all 24 SGP LRS families. SV statistics calculated using family-based multisample VCF file.**

#### Supplementary Figures

##### Supplementary Figure 1

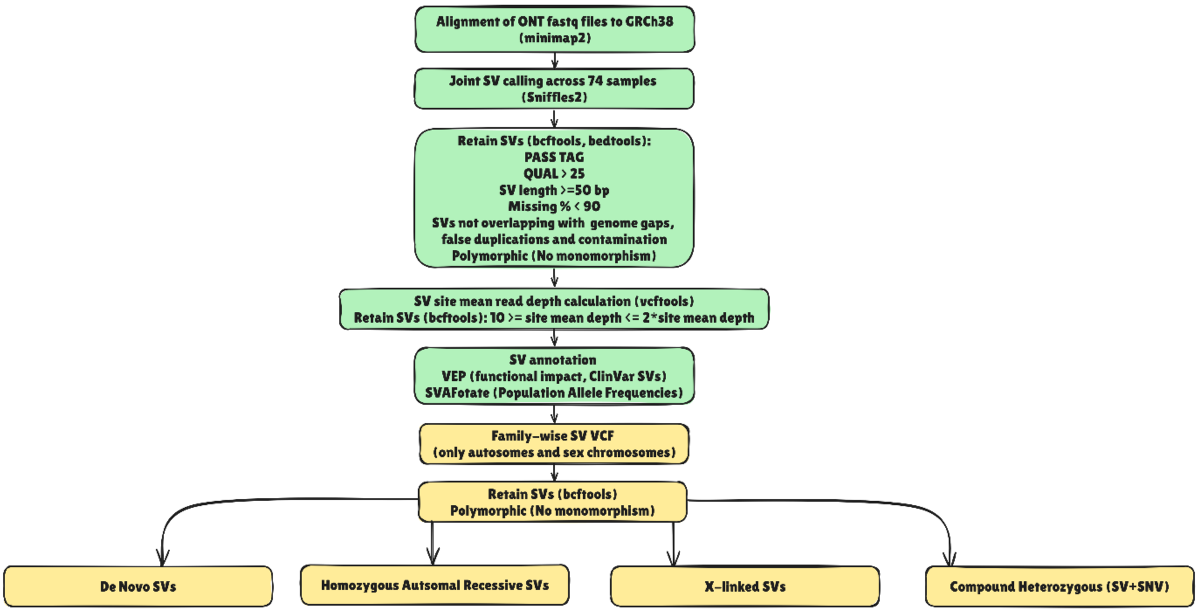


**Supplementary Figure 1: Schematic diagram representing the SV calling and filtering workflow. Green boxes represent bioinformatics steps for cohort-level SV calling and filtering. Yellow boxes represent bioinformatics steps applied to family-specific VCFs. Four modes of inheritance-based filtering were applied in the last step, as all parents were unaffected.**

#### Supplementary Figure 2

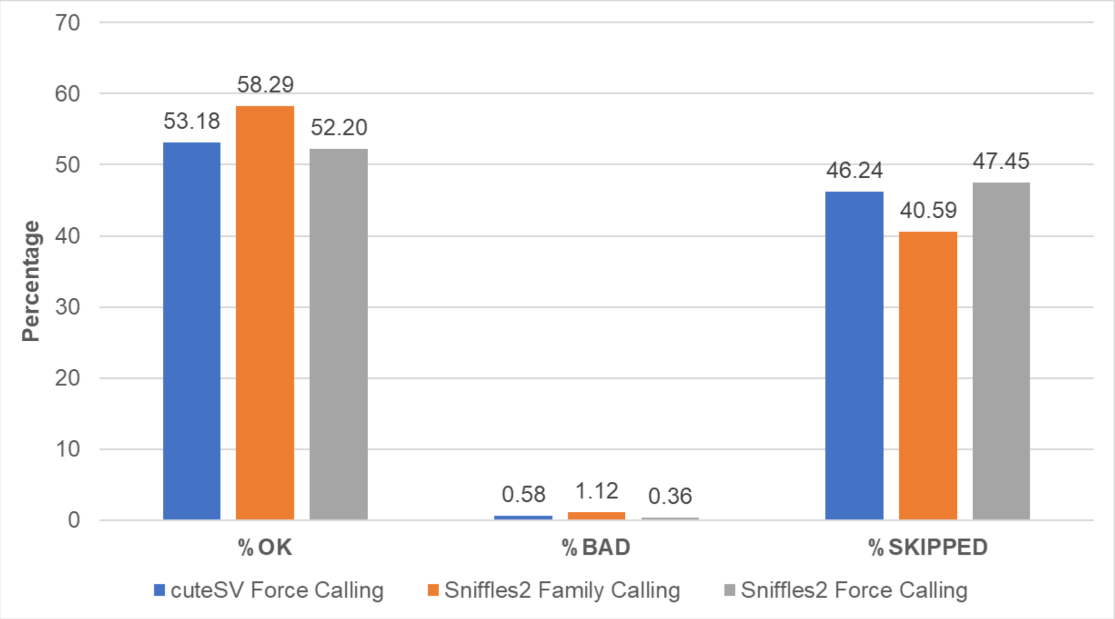


**Supplementary Figure 2: Mendelian consistency analysis plot showing %OK (% of genotypes at which the trio had no missingness and no Mendelian error), %BAD (% of genotypes at which the trio had a Mendelian error) and %SKIPPED (% of genotypes at which that trio had at least one individual missing and therefore could not be considered) across different multisample calling methodologies- cuteSV force calling, Sniffles2 family calling and Sniffles2 force calling. The trio family was randomly selected from the SGP LRS cohort for this analysis.**

#### Supplementary Figure 3

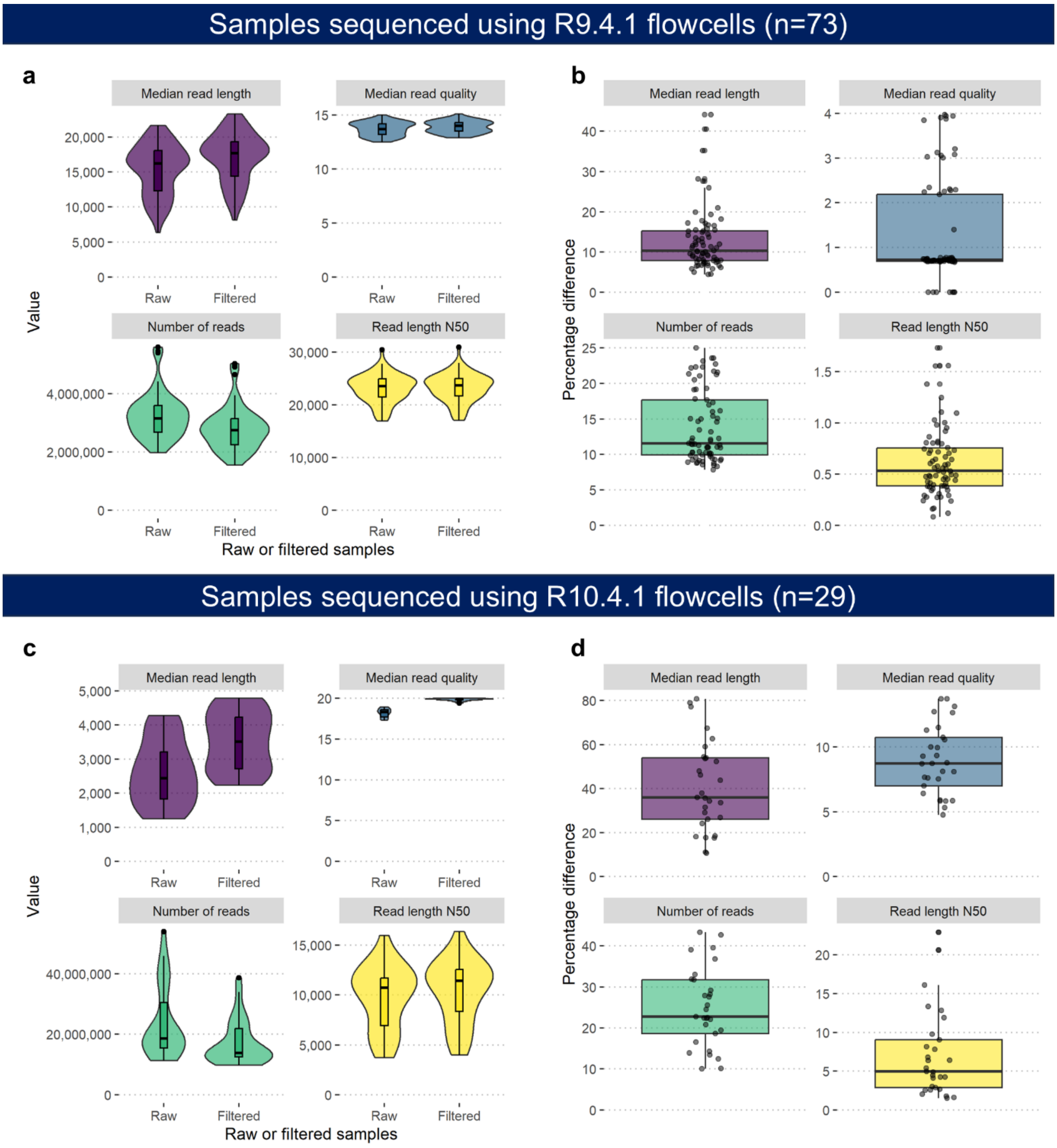


**Supplementary Figure 3: Comparison of QC metrics between raw and filtered samples sequenced using the R9.4.1 (subfigures a, b) and R10.4.1 (subfigures c, d) flowcells. (a, b) Median read length, median read quality, number of reads, and read length N50 are presented using violin plots with embedded boxplots for both raw and filtered samples. (c, d) Percentage difference between raw and filtered values for each QC metric, shown using boxplots with overlaid jittered data points.**

#### Supplementary Figure 4

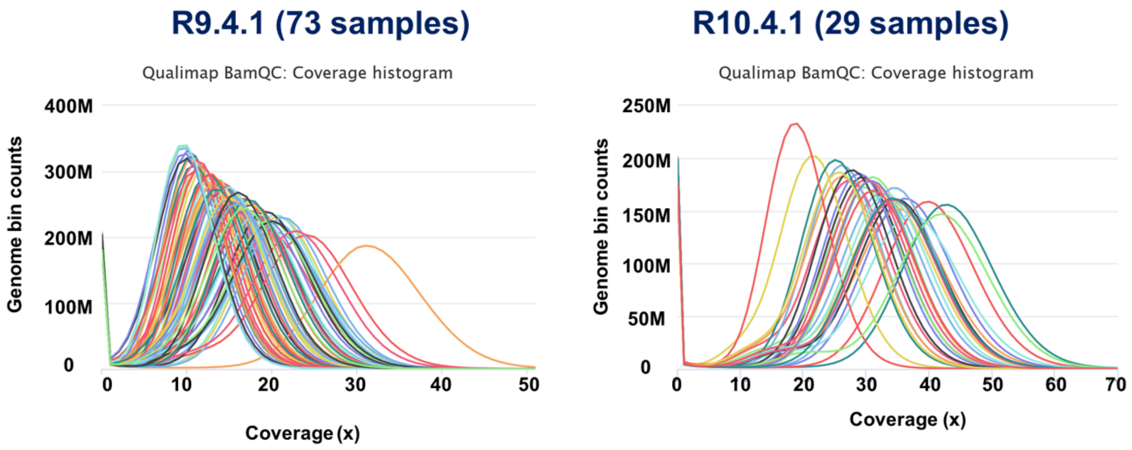


**Supplementary Figure 4: Depth of Coverage plots for samples sequenced using only R9.4.1 flowcells (left) vs R10.4.1 flowcells (right) calculated using Qualimap and plotted using MultiQC. Samples on the right show average coverage ranging from approximately 19x to 45x, whereas samples on the left show an average coverage which ranges from about 10x to 31x.**

**Supplementary Figure 5**

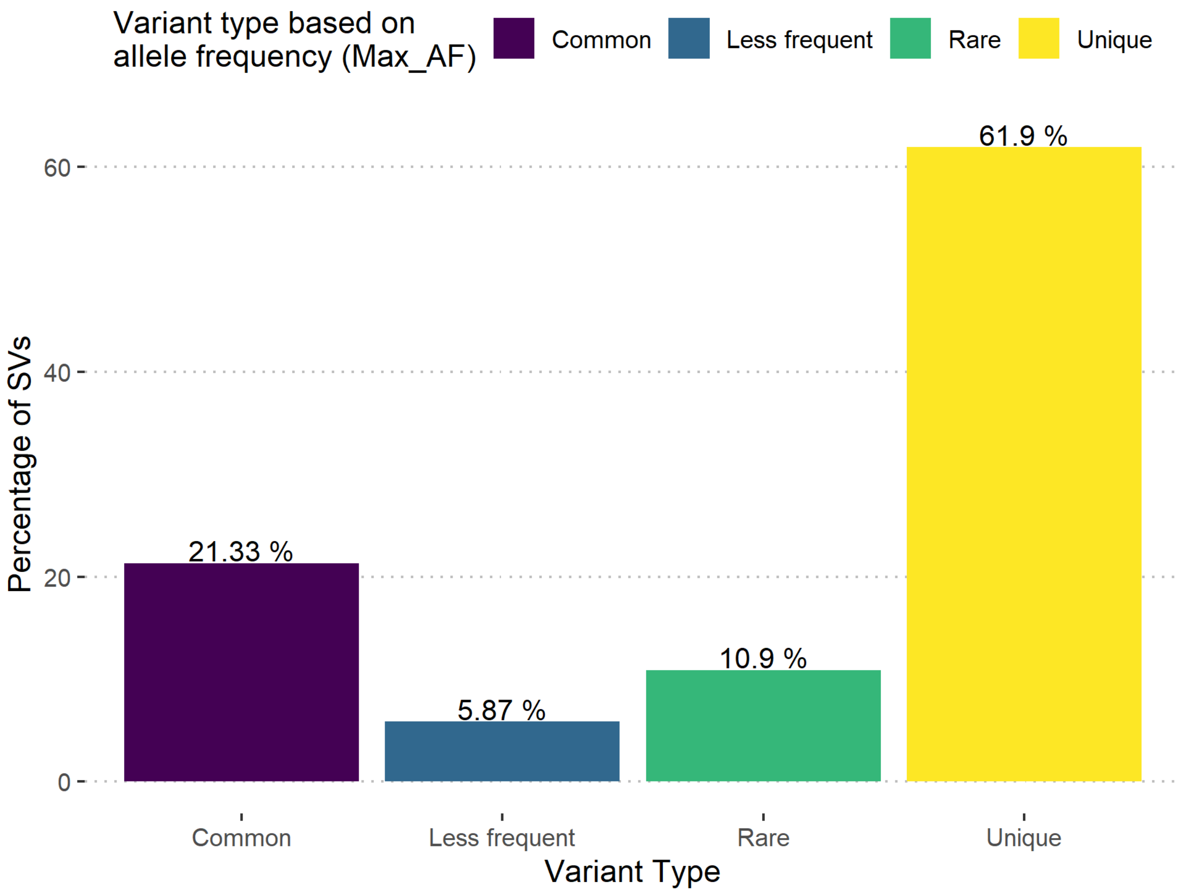


**Supplementary Figure 5: Barplot showing percentage of SVs in SGP LRS cohort stratified by variant type based on maximum allele frequency (Max_AF) from SVAFotate.**

#### Supplementary Figure 6

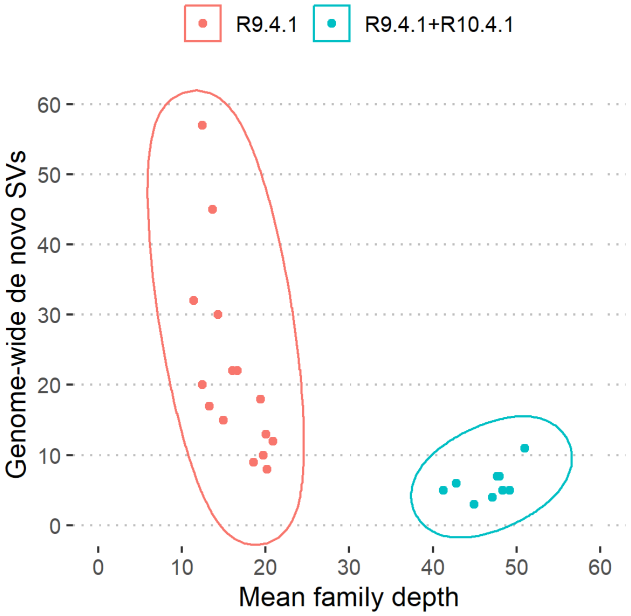


**Supplementary Figure 6: Number of genome-wide de novo SVs vs. mean family depth. Each dot represents a SGP LRS family, and the different colours tell if the samples from that family have been sequenced using just R9.4.1 flowcell (red) or using both R9.4.1 and R10.4.1 flowcells (blue). The two clusters signify that families sequenced using both flowcells led to a higher depth of coverage and a relatively lower number of de novo SVs genome-wide than families sequenced using just the R9.4.1 flowcell.**

#### Supplementary Figure 7

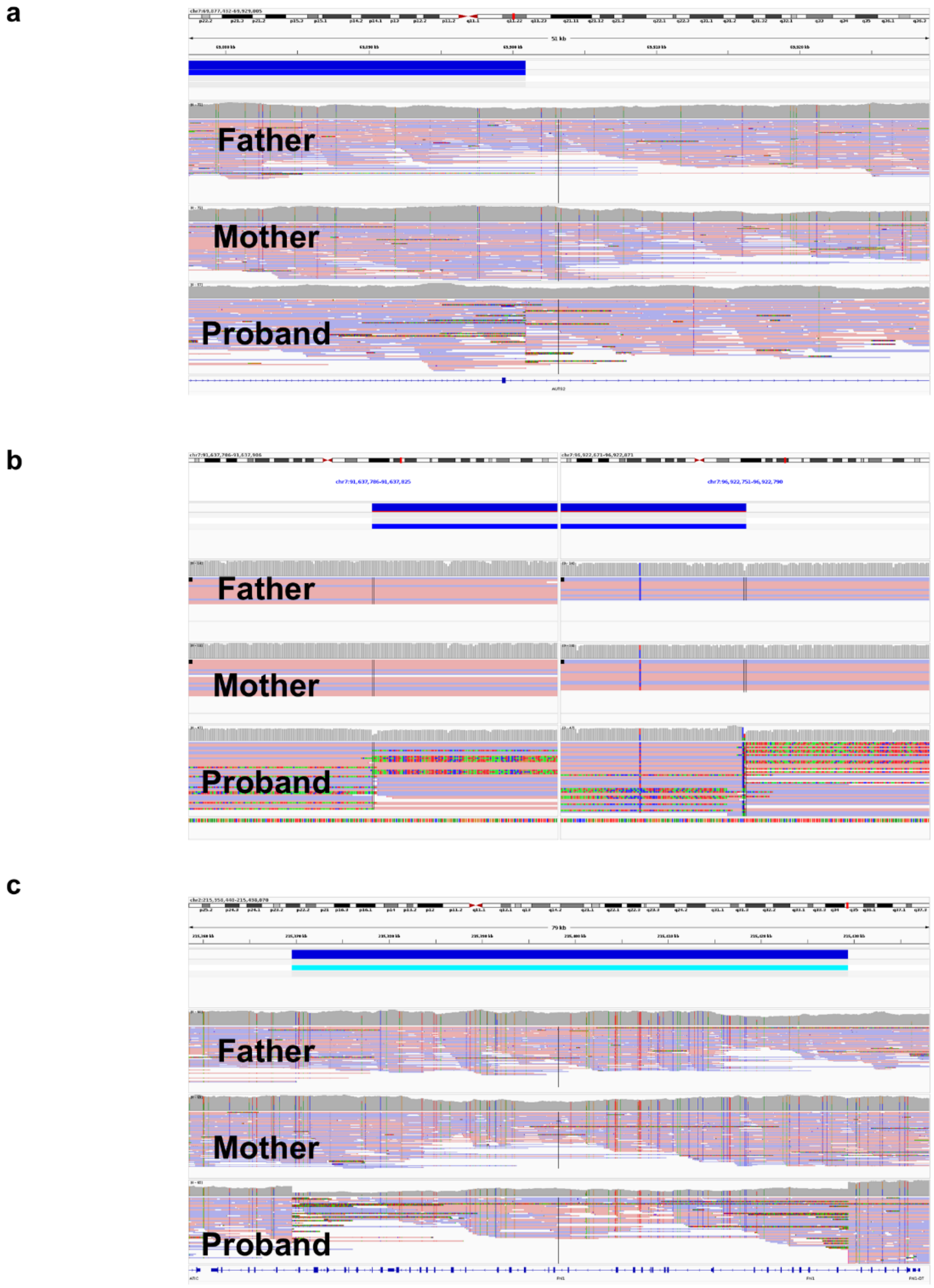


**Supplementary Figure 7: SGP LRS IGV plots showing *de novo* SV events in three exemplar families (all plots based on GRCh38 coordinates and sequenced by LRS). (a) One breakpoint of the *de novo* inversion in the proband from exemplar family 1 located in intron 2, disrupting *AUTS2*. (b) *De novo* inversion in the exemplar family 2 proband with breakpoints in two different regions in the non-coding part of chromosome 7. This IGV plot shows data only for the three family members (proband, mother, father) who were sequenced by LRS (c) Multiexon *de novo* heterozygous deletion of *FN1* in the proband from exemplar family 3 disrupting exons 6 to 41.**
